# Localised signalling networks co-opt TGF-β2 to promote an immuno-exclusive mesenchymal niche within human squamous cell carcinoma

**DOI:** 10.1101/2025.05.23.25327735

**Authors:** Matthew J Bottomley, Michael McKenna, Zehua Li, Noushin Zibandeh, Girishkumar Kumaran, Megat Abd Hamid, Elie Antoun, Eleni Ieremia, Lilian Brewer Lisboa, Trieu My Van, Tao Dong, Graham Ogg

## Abstract

Cutaneous squamous cell carcinoma (CSCC) is the most common metastasising malignancy affecting Caucasian populations. Advanced disease is associated with poor outcomes, with only half of patients responding to current immunotherapy regimens. Robust CD8+ T cell responses play a key role in determining CSCC outcome but understanding of local influences upon CD8+ behaviour within CSCC is limited. Here we show by multimodal tissue profiling alongside functional validation that TGF-β2 signalling in the setting of local inflammation, is co-opted to promote endothelial trans- differentiation into suppressive cancer-associated fibroblasts, representing a dominant mechanism by which malignant keratinocytes and fibroblasts regulate immune infiltration into fibrovascular niches at the leading tumour edge. Such niches demonstrate parameters predicting enhanced tumour invasion and immunotherapy resistance. Our results identify pathways which drive tumour invasiveness and exclude infiltrating effector lymphocytes at the tumour edge. We identify potential targets for therapy to target these niches and improve outcome in advanced disease.

## Introduction

Epidermal keratinocyte-derived cutaneous squamous cell carcinoma (CSCC) is the most common metastasising cancer in the Western world, diagnosed in over a million North American patients a year. Increasing annual incidence means CSCC causes twice as many cancer-specific deaths compared to melanoma each year [1, 2]. Risk factors include Caucasian ethnicity, increasing age, cumulative UV radiation exposure and immunodeficiency [3]. Excision is curative in early disease, though up to half of patients develop further asynchronous primary lesions [3, 4]. Metastatic disease, occurring in 4% of non-immunosuppressed cohorts, has historically been associated with a poor prognosis [5].

CSCC exhibits the highest mutational burden of any malignancy, particularly in the pre-malignant phase [6, 7], potentially creating multiple immunogenic neoantigens. Intact CD8+ T cell immunity determines CSCC development and outcome in pre-clinical models [8, 9]. Poor T cell tumour infiltration is associated with progression to advanced disease [10]. CSCC incidence and aggression is increased significantly in immunosuppressed populations such as organ transplant recipients [4].

Whilst immunotherapy has revolutionised outcomes in metastatic CSCC, less than half of patients demonstrate clinical response [11]. Pre-treatment tumour infiltrating lymphocyte (TIL) density in primary lesions predicts response to immunotherapy [12].

Previous studies of CSCC microenvironment heterogeneity have utilised pre-clinical models, low-plex antigen-specific staining, in vitro functional evaluation, and single-cell transcriptomic profiling [13–15]. Whilst of major value, these only provide limited resolution into the relevant influence of identified pathways upon localised TIL behaviour and particularly have been limited in advancing understanding of the contribution of non-immune cell populations. There is an unmet need to identify determinants of TIL accumulation in CSCC to develop novel therapeutic approaches and improve outcome in both immunocompetent (ICP) and immunosuppressed populations.

The advent of highly-multiplex, single cell resolution spatial profiling permits greater understanding of niche control within the TME and mechanisms underlying immune escape. In this study we leverage complementary spatial profiling approaches to investigate immunological heterogeneity in the TME of CSCC, identifying mechanisms associated with reduced TIL accumulation.

## Results

### Sample characteristics

CSCC, sampled from immunocompetent participants (ICP, n=6) and kidney transplant recipients (KTR, n=2), were submitted for histotranscriptomic analysis (Table 1). All participants were Caucasian [16]. CD8+ T cell density, evaluated by immunoenzymatic staining, was dramatically reduced in CSCC from KTR compared to ICP, consistent with previous reports [13].

**Table 1:**
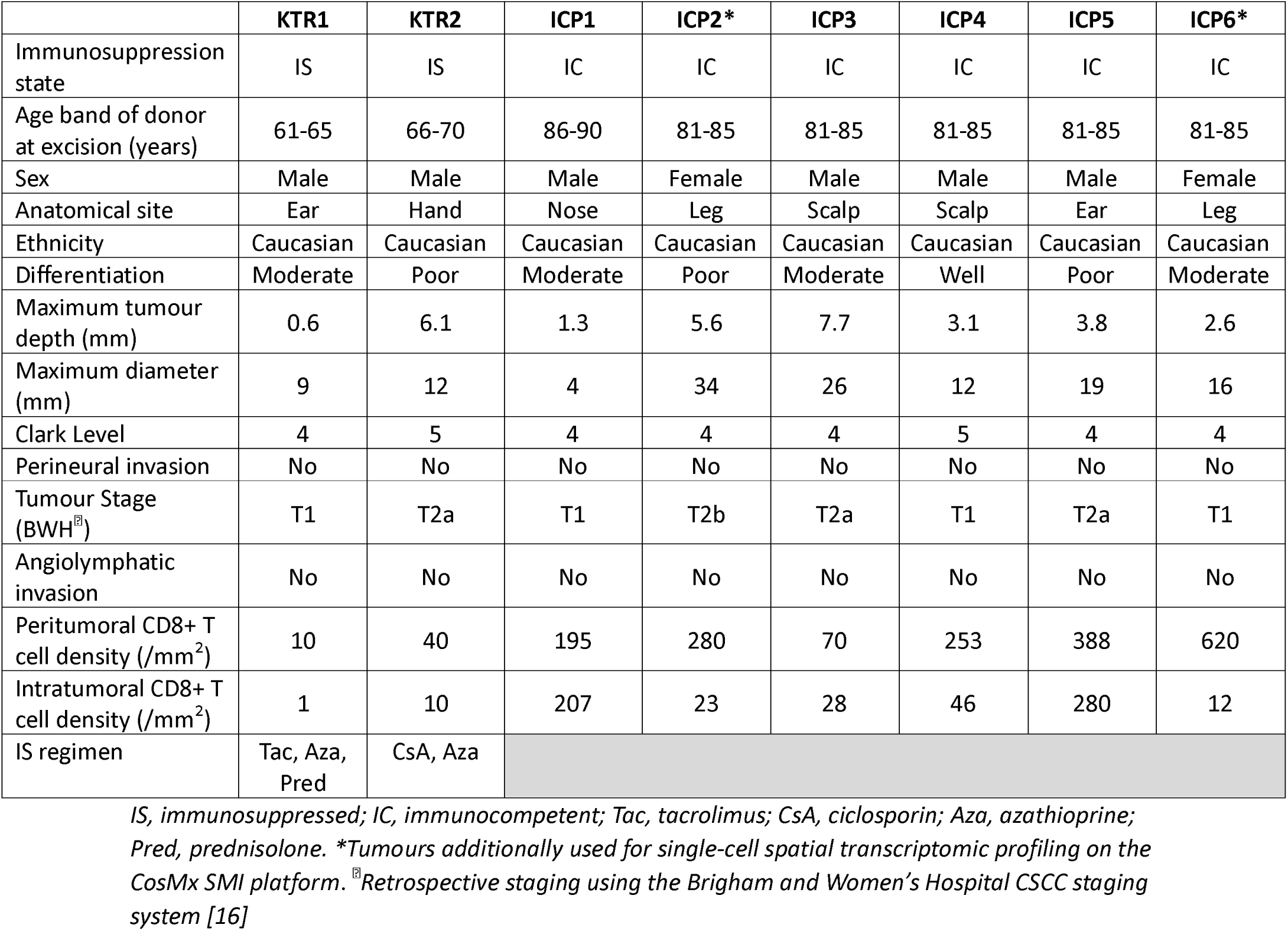
Clinical and histopathological features of tumours analysed by spatial profiling

|  | KTR1 | KTR2 | ICP1 | ICP2* | ICP3 | ICP4 | ICP5 | ICP6* |
| --- | --- | --- | --- | --- | --- | --- | --- | --- |
| Immunosuppression state | IS | IS | IC | IC | IC | IC | IC | IC |
| Age band of donor at excision (years) | 61-65 | 66-70 | 86-90 | 81-85 | 81-85 | 81-85 | 81-85 | 81-85 |
| Sex | Male | Male | Male | Female | Male | Male | Male | Female |
| Anatomical site | Ear | Hand | Nose | Leg | Scalp | Scalp | Ear | Leg |
| Ethnicity | Caucasian | Caucasian | Caucasian | Caucasian | Caucasian | Caucasian | Caucasian | Caucasian |
| Differentiation | Moderate | Poor | Moderate | Poor | Moderate | Well | Poor | Moderate |
| Maximum tumour depth (mm) | 0.6 | 6.1 | 1.3 | 5.6 | 7.7 | 3.1 | 3.8 | 2.6 |
| Maximum diameter (mm) | 9 | 12 | 4 | 34 | 26 | 12 | 19 | 16 |
| Clark Level | 4 | 5 | 4 | 4 | 4 | 5 | 4 | 4 |
| Perineural invasion | No | No | No | No | No | No | No | No |
| Tumour Stage (BWH<sup>®</sup>) | T1 | T2a | T1 | T2b | T2a | T1 | T2a | T1 |
| Angiolymphatic invasion | No | No | No | No | No | No | No | No |
| Peritumoral CD8+ T cell density (/mm<sup>2</sup>) | 10 | 40 | 195 | 280 | 70 | 253 | 388 | 620 |
| Intratumoral CD8+ T cell density (/mm<sup>2</sup>) | 1 | 10 | 207 | 23 | 28 | 46 | 280 | 12 |
| IS regimen | Tac, Aza, Pred | CsA, Aza |  |  |  |  |  |  |

IS, immunosuppressed; IC, immunocompetent; Tac, tacrolimus; CsA, ciclosporin; Aza, azathioprine; Pred, prednisolone. \*Tumours additionally used for single-cell spatial transcriptomic profiling on the CosMx SMI platform. <sup>®</sup>Retrospective staging using the Brigham and Women's Hospital CSCC staging system [16]

### Area of interest (AOI) sampling

77 AOI (median AOI per tumour: 10 (range 5 – 17)) were chosen for histotranscriptomic profiling (Fig. 1a-c and S1-S2) based on (a) histological appearance and pan-cytokeratin (panCK) positivity, indicating malignant keratinocyte morphology (tumour-rich regions, TRR, n=32) and (b) clustering of CD8-staining cells in the panCK-negative stroma (immune-rich regions, IRR, n=37). One additional AOI per section was taken from adjacent morphologically normal skin (perilesional normal skin, PNS, n=8).

**Figure 1:**
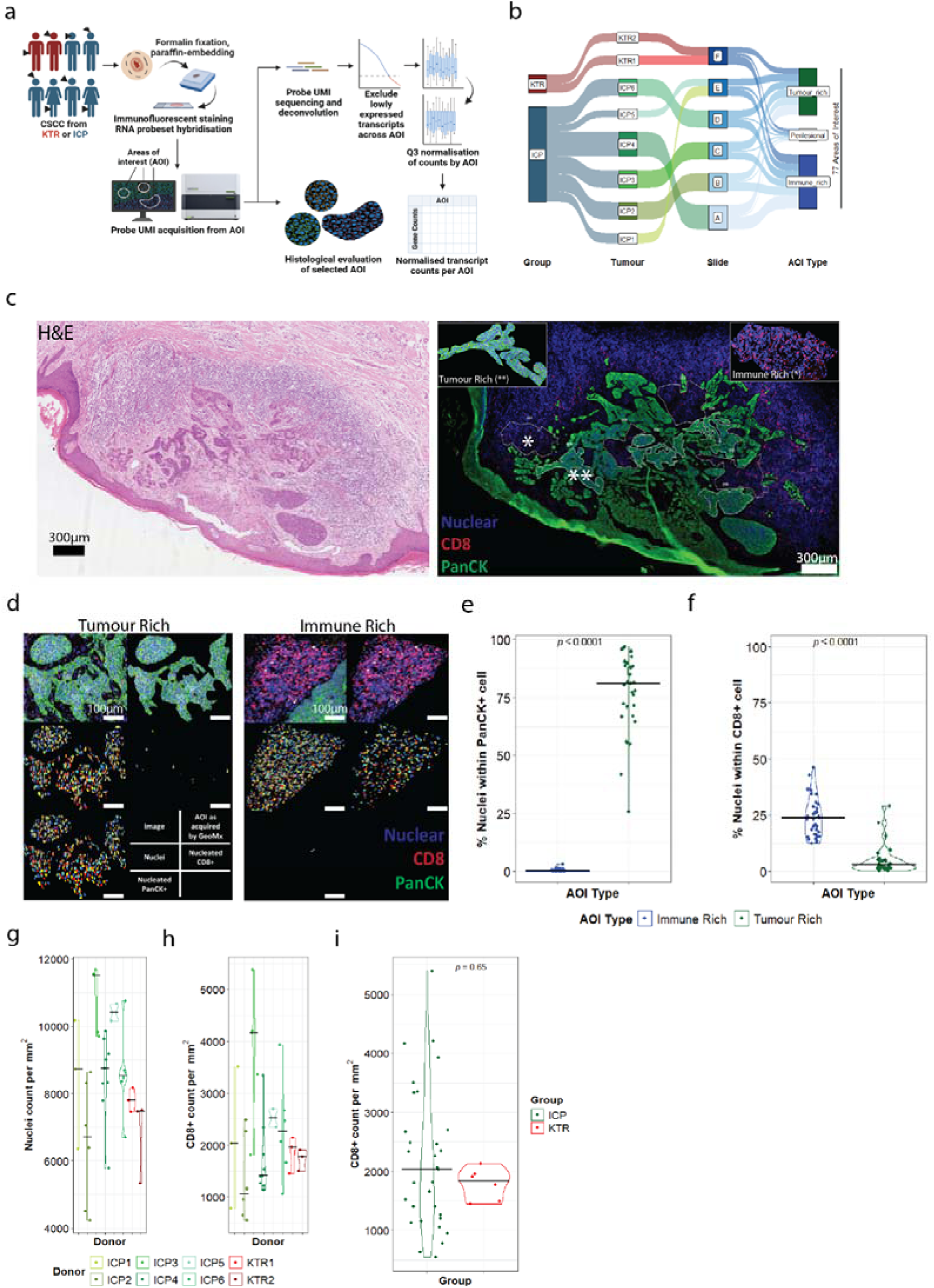
Experimental design and histological analysis of CSCC areas of interest (AOI) (A) Workflow for data acquisition, QC and histological evaluation. Black triangles indicate site of excision. (B) Sankey diagram indicating GeoMx experimental design. CSCC from KTR (n=2 donors) and ICP (n=6 donors) were mounted onto slides (dual-mounted where possible), with IRR and TRR identified based on morphology and PanCK and CD8 staining. (C) Example Haematoxylin & Eosin (H&E) image (left) and corresponding CSCC image (right) used for AOI acquisition using the Bruker Spatial Biology GeoMx™ DSP platform. Probes within AOI were acquired from these slides with magnified inset examples of each region type. Blue represents SYTO 13 (nuclear) staining; Green pan-cytokeratin staining; red CD8A staining. (D) AOI were evaluated histologically using a CellProfiler pipeline to identify and enumerate nucleated cells expressing PanCK and CD8 within each AOI. Analysis of immune rich and tumour rich AOI, with (L-R by row) IF image; IF image after application of mask to create AOI as acquired by GeoMx; detected nuclei; nucleated CD8+ cells; nucleated PanCK+ cells. Blue represents SYTO 13 (nuclear) staining; Green pan-cytokeratin staining; red CD8A staining. White bars denote scale (100µm in all cases) Proportion of total nucleated cells staining for PanCK (E) and CD8 (F) within Immune and Tumour Rich Regions. Nuclear (G) and CD8+ density (H) within IRR, stratified by donor (G-H) and immunosuppression status (I). Comparison of distribution between groups was tested by Kruskal-Wallis test. AOI; area of interest. CSCC; cutaneous squamous cell carcinoma. ICP; immunocompetent. KTR; kidney transplant recipient.

### Morphotranscriptomic evaluation of CSCC AOI

AOI selection was validated by enumeration of panCK and CD8 staining cells within TRR and IRR (Fig. 1d-f). Parameters obtained by CellProfiler analysis of AOI correlated strongly with those calculated by GeoMx automated quantification and inferred cell abundance (Extended Data Fig. 1a-c). IRR AOI demonstrated marked intra- and inter-tumoral heterogeneity in both absolute nuclear and CD8+ density (Fig. 1g-h) and relative proportion amongst nucleated cells within IRR AOI (Extended Data Fig. 1d), which was not influenced by immunosuppression state (p=0.65 by Kruskal Wallis test, Fig. 1i and Extended Data Fig. 1e).

After quality control, dimensional reduction of 12,935 normalised gene counts, detected above the limit of quantification (Extended Data Fig. 2), indicated that the AOI type was the primary contributor to transcriptomic variation as expected (Fig. 2a). Notably, immunosuppression status was not a major independent contributor (Fig. 2b and Extended Data Fig. 3). Genes showing the greatest variation across all AOI related to keratinocyte function (CALML5, FLG, LCE1A/3D, IVL, CASP14, SPRR1A/1B/2B/2E and SERPINB13), immunoglobulin production (IGLL5, IGHA1, JCHAIN), innate immunity (LCN2, DEFB4B) and extracellular matrix remodelling (MMP7, KLK6, Fig. 2b). Following linear modelling, 3,103 genes were differentially expressed between IRR and TRR (1.5 absolute-fold change and adjusted p value <0.05, Fig. 2c and Supplemental Data).

**Figure 2:**
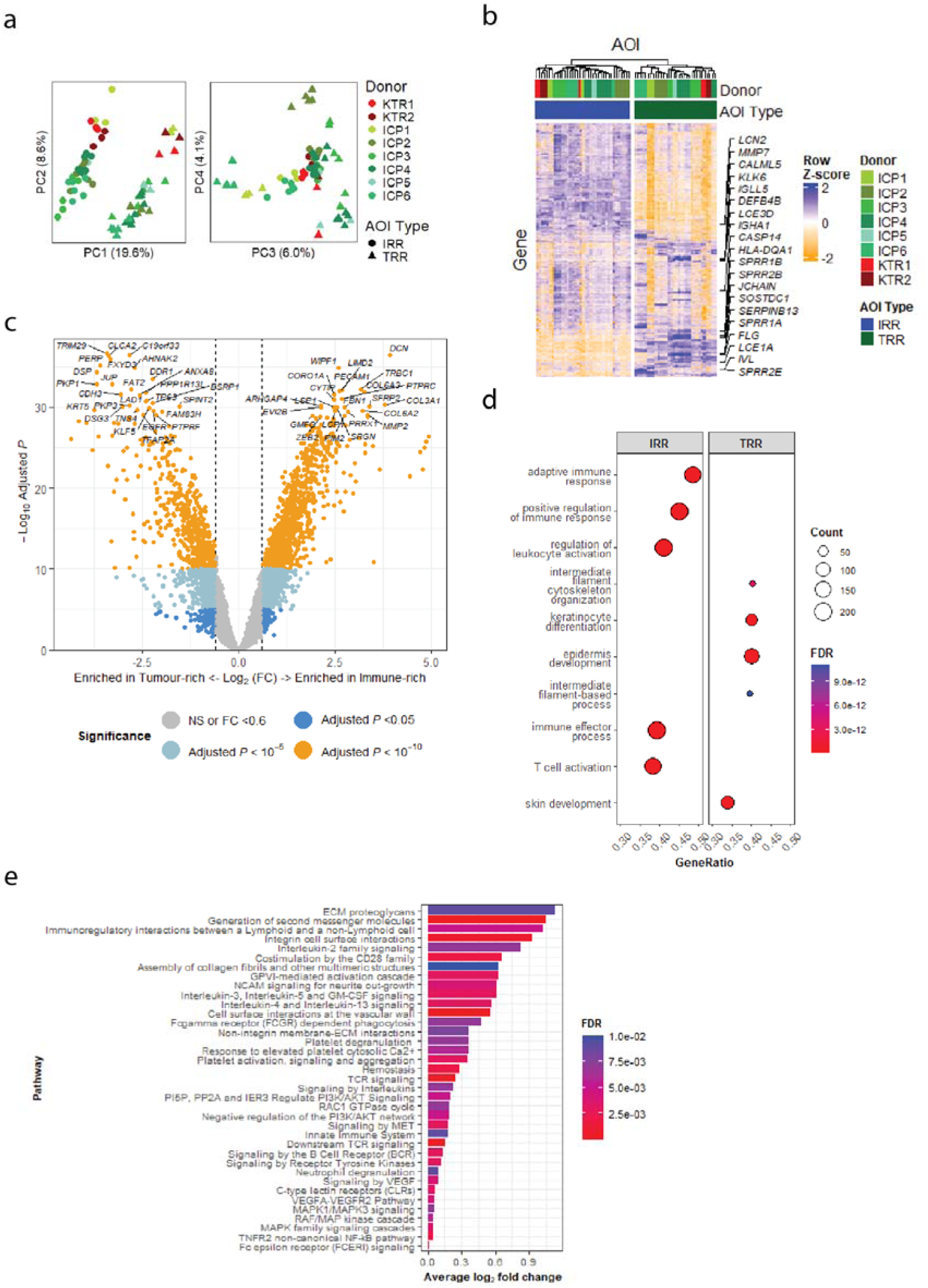
Differential gene expression and pathway analysis in immune- and tumour-rich AOI. (A) Principal component analysis plot demonstrating first four principal components of gene expression, coloured by donor ID (KTR coloured in red, ICP in green). (B) Heatmap of the top 20% most variable genes, annotated by donor ID and AOI type and ordered by unsupervised hierarchical clustering. The top 20 most variable genes across AOI are annotated. (C) Volcano plot illustrating differentially expressed genes using ‘differential expression for repeated measures’ (‘dream’) approach to linear modelling between immune-rich and tumour-rich regions (IRR and TRR, respectively). The top 50 differentially expressed genes are highlighted. (D) Top five differentially enriched gene sets from Gene Ontology ‘Biological Processes’ subontology between IRR and TRR. (E) Reactome pathways significantly enriched in IRR using PADOG. ‘PADOG’, ‘Pathway Analysis with Downweighting of Overlapping Genes’. Only pathways with an FDR <0.05 and >30 genes present are shown. ‘KTR’, kidney transplant recipient; ‘ICP’, immunocompetent. ‘AOI’, area of interest. ‘FDR’, false discovery rate.

Gene sets relating to immunity were upregulated in IRR, whilst those relating to keratinisation and epithelial function were upregulated in TRR (Fig. 2d and Extended Data Fig. 4a-b). Pathway analysis revealed upregulation of pathways relating to innate, T cell and humoral immune response and cytokine and chemokine signalling in IRR. There was notable enrichment of pathways indirectly related to immune function, including haemostasis, extracellular matrix remodelling and neuronal activity (Fig. 2e), many of which were enriched to a greater degree than immunological pathways. These data reveal the expected distinct transcriptomic patterns between TRR and IRR and further highlight the significant activity amongst indirect immunological processes in the tumour microenvironment local to CD8+ T cell aggregation.

### Cellular deconvolution

Cellular deconvolution was used to infer the presence of cell types within AOI (Fig. 3a) [18]. A reference matrix of gene expression profiles for 34 cell populations was constructed utilising a recently-published single-cell dataset for CSCC (Fig. 3b) [15]. This reference matrix was used to estimate the abundance of cell populations within CSCC AOI.

**Figure 3:**
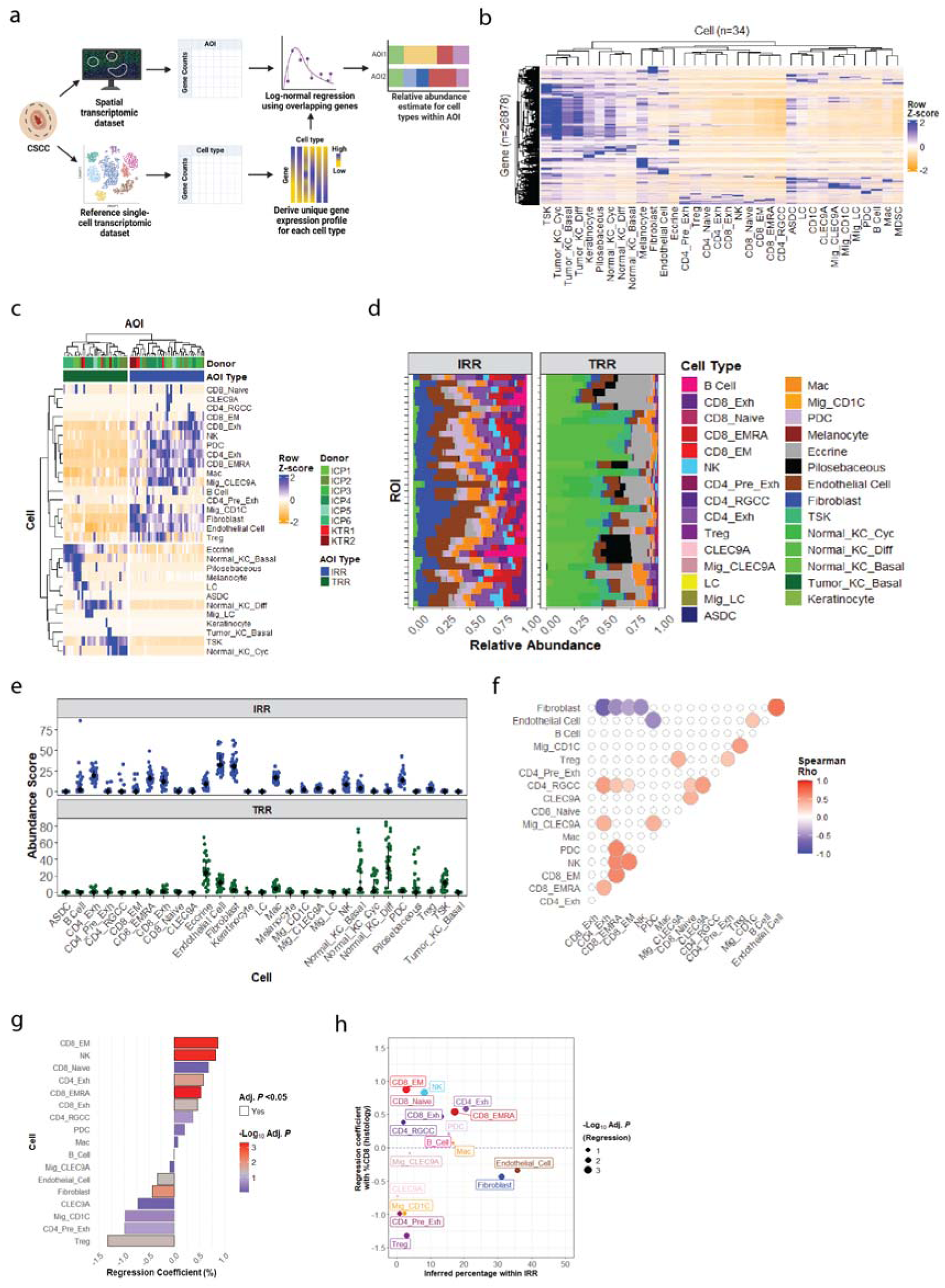
Cellular deconvolution of cell populations. (A) Summary of cellular deconvolution workflow. Abundance of cell populations within AOI were inferred by log-normal regression using reference single-cell gene expression profiles. (B) Heatmap of reference transcriptomic profile for each cell type, derived from [15] (C) Heatmap and unsupervised clustering of deconvoluted cell abundances by AOI, after removal of undetectable cell populations. (D) Stacked bar chart showing relative inferred abundance of cell types within IRR and TRR. (E) Inferred abundance by cell type within IRR and TRR. Median and IQR of inferred abundance are shown for each population. (F) Correlogram of deconvoluted population abundance across IRR: only correlations significant by Spearman rank test, after adjustment for repeated testing, are shown. (G) Linear modelling of inferred cell abundance and histologically determined CD8+ percentage of nucleated cells across IRR AOI, controlling for donor. Bar colour indicates adjusted P value (low = red, high = blue) whilst black outline on bars indicates those with adjusted P <0.05. (H) Scatterplot of mean inferred cell type abundance across all IRR and association with histological CD8+ T cell accumulation. CD8_EM = CD8+ Effector Memory; NK = Natural Killer; CD4_Exh = Exhausted CD4+ T cell; CD8_EMRA = CD8+ Effector Memory CD45RA+; CD8_Exh = Exhausted CD8+ T cell; CD4_RGCC = RGCC-expressing CD4+ T cell; PDC = plasmacytoid dendritic cell; Mac = Macrophage; Mig_CLEC9A = Migrating CLEC9A+ dendritic cell; CLEC9A = CLEC9A+ dendritic cell; Mig_CD1C = Migrating CD1C+ dendritic cell; CD4_Pre_Exh = Pre- exhausted CD4+ T cell; Treg = Regulatory T cell.

29 of 34 cell populations were inferred as present, with abundance profile forming two distinct clusters consistent with AOI Type. Keratinocyte, melanocyte and eccrine populations were enriched within TRR, whilst IRR were characterised by accumulation of T cell, B cell and myeloid populations (Fig. 3c).

Dendritic cells were inferred to form distinct clusters across compartments, with Langerhans and AXL+SIGLEC6+ dendritic cells (ASDC) present in TRR but not IRR, whilst plasmacytoid (pDC) and CLEC9A+ dendritic cells were found predominantly in the IRR. Endothelial and fibroblast populations were inferred to be the most frequent cell populations within IRR (Fig. 3d-e). Correlating the inferred abundance of IRR cell populations across AOI revealed the co-abundance of endothelial cells and fibroblasts, which negatively correlated with the abundance of effector cytolytic populations such as NK cells, CD8+ EM and CD8+ EMRA (Fig. 3f). Effector cytolytic populations were co-abundant and were positively correlated with the presence of pDC and CD4+RGCC+ populations. Using linear modelling to account for repeated donor sampling, increased CD8+ T cell proportion and absolute density within IRR (determined histologically) correlated with aggregation of other effector cells, while regions rich in fibroblasts, endothelial cells, and Tregs exhibited reduced CD8+ T cell presence (Fig. 3g and Extended Data Fig. 4c). Combining these analyses, cell-specific influences upon CD8+ accumulation within CSCC TME can be delineated (Fig. 3h). Treg represent a dominant, albeit relatively infrequent, suppressor of CD8+ T cell accumulation within the TME. Conversely, endothelial and fibroblastic (fibrovascular) populations demonstrate a weaker negative association with CD8+ T cell accumulation but together account for two-thirds of the TME population (Fig. 3h).

### Tumour-specific transcriptomic modules drive phenotypic variation within CSCC TME

To explore drivers of phenotypic variation across IRR in an unbiased fashion, pathway and deconvolution data were incorporated with unsupervised weighted co-expression gene network analysis (WGCNA) [19]. 19 distinct modules of gene co-expression were identified within IRR (Extended Data Fig. 5c and Supplemental Data). Modules demonstrated distinct correlation to inferred cell populations (Fig. 4a) and gene set enrichment profiles (Fig. 4b, Extended Data Fig. 5d and Supplemental Data), facilitating annotation of 13 gene modules (Extended Data Fig. 5e). Notably, those relating to keratinisation, extracellular matrix (ECM) formation and remodelling and angiogenesis (hereafter termed ‘fibrovascular modules’) had a strong negative unadjusted correlation with inferred effector T cell abundance, with the opposite seen for the T cell response (purple) module (Fig. 4c).

**Figure 4:**
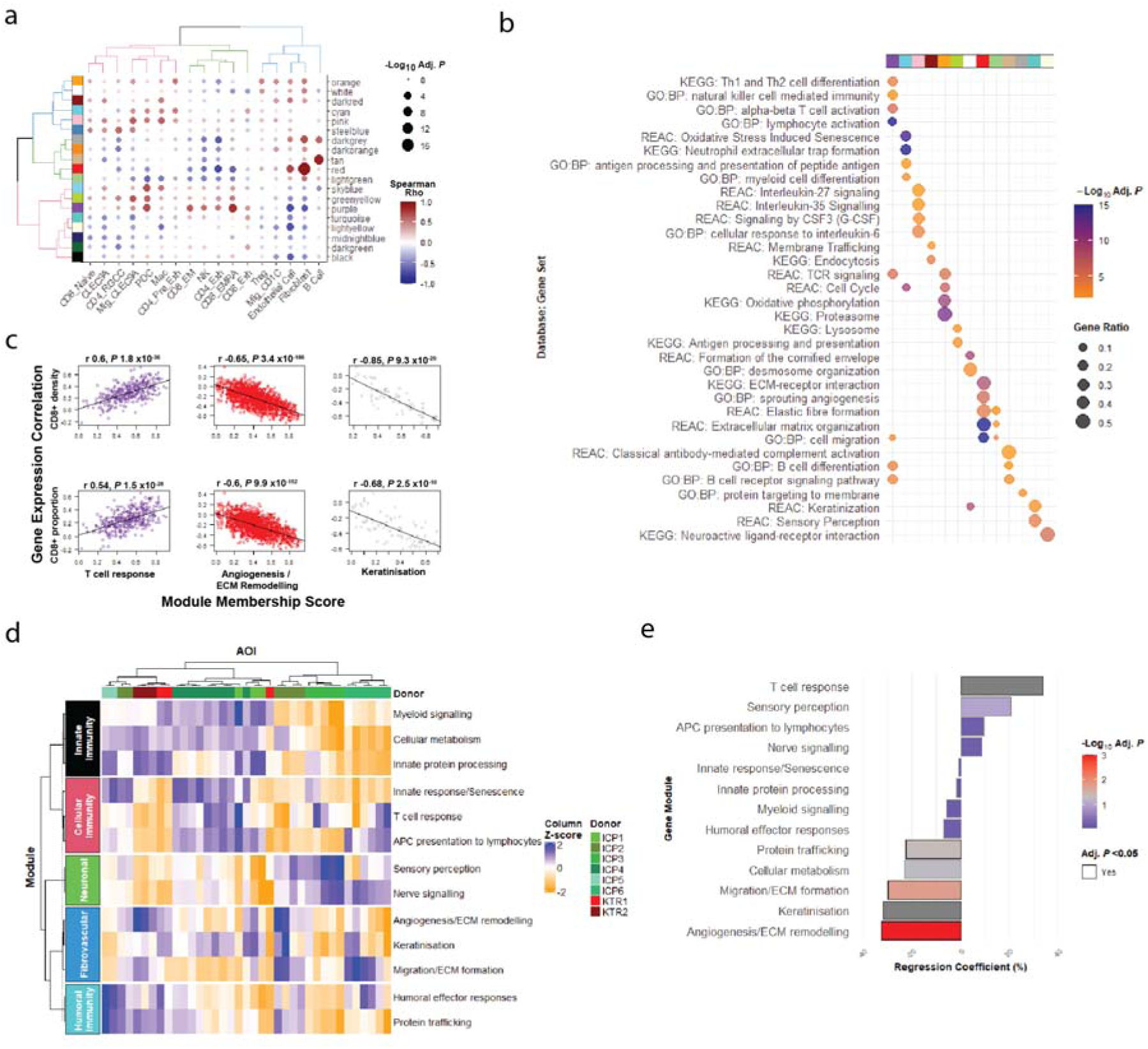
Gene network construction and association with CD8+ T cell accumulation. (A) Correlation plot of gene modules with inferred cell abundance. Points are sized by adjusted p value and coloured by Spearman rho correlation (blue = negative correlation; red = positive correlation). Module colours are shown on the left axis, for comparison with Fig. 4B. (B) Functional gene set enrichment within modules; annotated names are provided on the top y-axis, with module colour (corresponding to those in Fig. 4A) on the bottom y-axis. Point size corresponds to gene ratio and colour corresponds to-Log_10_ adjusted P value. Only modules with enriched gene sets are shown. (C) Spearman rank correlation of individual gene module membership with corresponding correlation with histologically- determined CD8+ T cell density (top row) and proportion (bottom row) for selected modules. Rho statistic and P values are provided above each plot. (D) Heatmap demonstrating module activity (based on AOI eigenvalue) and clustering across IRR AOI. Based on clustering, ‘supermodules’ of related modules are annotated (light blue = Humoral Immunity; dark blue = Fibrovascular; Green = Neuronal; Red = Cellular Immunity; Green = Innate Immunity (E) Linear modelling of module eigengene value and histologically-determined CD8+ T cell percentage (left) across IRR AOI, adjusted for donor. Bar colour indicates adjusted P value (low = red, high = blue) whilst black outline on bars indicates those with adjusted P <0.05.

Evaluation of individual module activity demonstrated distinct patterns of enrichment within AOI, permitted identification of supermodules relating to innate, cellular and humoral immunity, and fibrovascular/neuronal activity (Fig. 4d). Although individual AOI demonstrated distinct patterns of module enrichment, there was clustering by tumour, suggesting patterns of module activity are distinct between tumours.

Cross-validation with histological data confirmed CD8+ T cell accumulation demonstrated a positive correlation with module membership of genes within the T cell response network, and to a lesser extent the other modules within the cellular immunity supermodule (Fig. 4e and Extended Data Fig. 5f). There was a strong negative correlation with fibrovascular module activity.

Taken together, these results illustrate that fibrovascular populations are co-localised in the tumour stroma, consistent with previous reports [15]. These create fibrovascular TME niches where accumulation of CD8+ T cells and other effector populations is dampened.

### Signalling pathways within IRR

Having established a role for fibrovascular networks in dampening CD8+ T cell accumulation within the TME, the ligand-receptor interactions (LRI) and associated signalling pathways which may be driving this effect were evaluated. IRR were evaluated independently of TRR, given the distinctive cellular and transcriptomic features, leading to LRI heterogeneity (Extended Data Fig. 6A). 144 ligands and 98 receptors within IRR, with at least one moderate L-R expression correlation and significant downstream pathway association were analysed (Fig. 5a-b and Supplemental Data). No ligands were identified within the keratinisation, protein trafficking or innate protein processing modules. Distinct relationships between ligands from different modules and CD8 T cell accumulation were observed (Fig. 5c and Extended Data Fig. 6b); most ligands had an inhibitory association with CD8 accumulation. Ligands with highest membership of their respective module and strongest negative association with CD8 accumulation were notably located within the fibrovascular modules. Many of these genes related to ECM production, such as collagen and glycosaminoglycans (Fig. 5c inset). After linear modelling, the strongest negative association with CD8 accumulation was unexpectedly seen for the interaction between TGFB2 and ENG, encoding transforming growth factor beta 2 (TGF-β2) and endoglin respectively (Fig. 5d).

**Figure 5:**
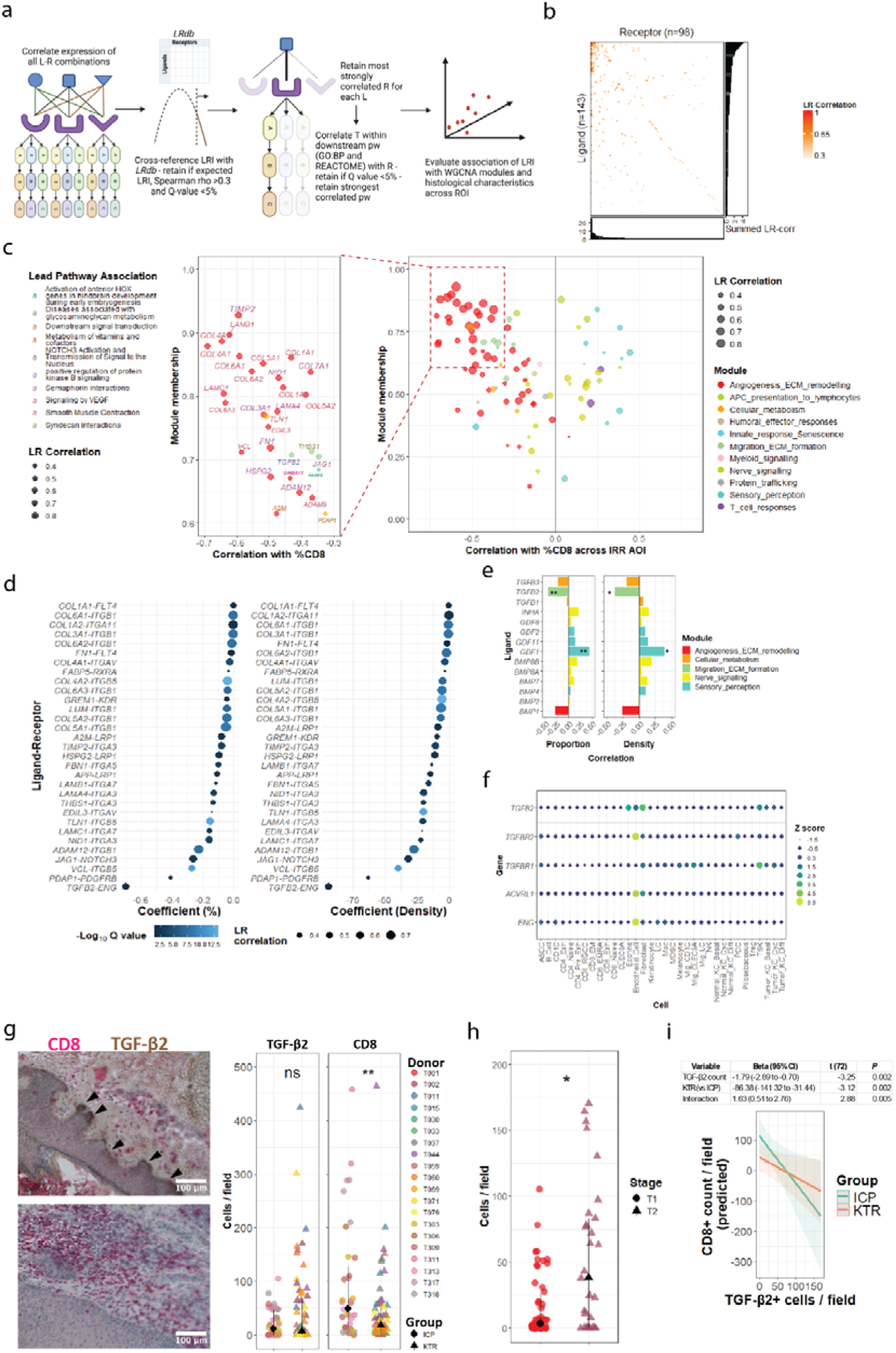
Ligand-receptor interactions within IRR AOI. (A) BulkSignalR workflow, showing how ligand (L), receptor (R) and pathways (pw) triples are created based on L-R-target (T) correlations. (B) Heatmap of all ligands and receptors with at least one significant correlation, prior to LR and pw collapse. (C) Scatterplot of ligand correlation with histologically-determined CD8+ T cell density across AOI, and module membership. Inset on left highlights the lead pathway association for those ligands with the strongest negative correlation with CD8 density. (D) Linear modelling of lead inhibitory ligands for CD8 accumulation. (E) Correlation of TGF-β2 superfamily gene expression with histologically determined CD8 infiltration within IRR AOI. * p<0.05, **p<0.01 by Spearman rank test. (F) Expression of TGFB2 and its ligands upon cell types within Ji et al dataset . (G) Dual-colour immunohistochemistry for CD8 (pink) and TGF-β2 (brown) – example staining of a field with high TGF- β2 and low CD8 staining (top-left) and low TGF-β2 and high CD8 staining (bottom-right). Black triangles highlight areas of TGF-β2 staining at the leading tumour edge. Right: Positively stained TGF- β2 (left) and CD8 (right) cells per field stratified by group (circles = NIC, triangles = KTR) ns not significant, **p<0.01 by Mann-Whitney test. (H) TGF-β2 stained cells per field, stratified by BWH histological staging. Red circles denoteT1, brown triangles denote T2, with a black circle/triangle indicating the arithmetic mean for each group. P <0.05 by Mann-Whitney test. (I) Mixed effect modelling for CD8+ count by TGF-β2+ count per field, incorporating study group (KTR or ICP) and repeated donor sampling. Top: results of mixed effect modelling. Bottom: predicted CD8+ count by TGF-β2+ count, stratified by group. Orange line indicates KTR (orange shading 95% confidence intervals); Green line indicates ICP (green shading represents 95% confidence intervals).

The TGF superfamily are promiscuous in terms in receptor binding, and so receptor-downstream pathway analysis was next undertaken. To confirm this was a TGFB2-ENG specific interaction effect associated with diminished CD8 accumulation, all alternative ligands and receptors present within the superfamily were evaluated for their association with CD8 accumulation (Fig. 5e and Extended Data Fig. 6c). TGFBR3 (betaglycan) and ACVRL1 (ALK1) were also included in this analysis as these have been demonstrated to modulate downstream signalling [20]. ENG and ACVRL1 demonstrated a significant association with diminished CD8 accumulation. As the ENG-ALK1 complex may also bind other ligands, including BMP-9 (GDF2), reverse validation was undertaken for alternative ligands for ENG. TGFB2 was the only ligand demonstrating this association (Fig. 5e). To evaluate potential sources of TGF-β production and signalling, gene sets relating to TGF-β were interrogated for their enrichment within modules. Gene sets relating to TGF-β production were found solely within the ‘ECM formation’ module, whilst TGF-β response-related sets were found solely within the Angiogenesis/ECM remodelling’ module (Extended Data Fig. 6d). Cross-validation, using the reference scRNAseq dataset previously utilised for cellular deconvolution [15], confirmed TGFB2 expression was highest in fibroblast, eccrine and tumour-specific keratinocyte (TSK) populations, whilst ENG and ACVRL1 expression was highest in endothelial cells (Fig. 5f). Providing further validation, the previously identified TGFB-related gene set enrichment scores were highest in a hierarchical cluster containing basal and tumour keratinocyte, endothelial and fibroblast populations (Extended Data Fig. 6e).

To confirm the inverse relationship between TGF-β2 and CD8 expression at protein level, 20 CSCC (including the 8 evaluated by GeoMx DSP) were co-stained for TGF-β2 and CD8. TGF-β2 expression was predominantly found at the leading tumour edge, with a trend towards higher TGF-β2 expression, and a significant reduction in CD8 infiltration, in KTR compared to ICP (Fig. 5g). TGF-β2 expression was increased in tumours exhibiting histological high-risk features for metastasis (Fig 5h). An inverse association was found between TGF-β2 expression and local CD8+ T cell infiltration irrespective of whether CSCC were from immunocompetent or immunosuppressed individuals (Fig. 5i), though the relationship was blunted in KTR.

The tumour inflammation score (TIS) is a validated bulk gene expression signature, with increased score predictive of clinical response to immunotherapy across multiple cancer types. We hypothesised that fibrovascular areas of CSCC would exhibit reduced local TIS. We evaluated individual components and their association with TGFB2 expression [21]. 15 of 18 genes within the TIS were downregulated in TGFB2 enriched areas, with 7 achieving statistical significance (P <0.05 by Spearman rank test, Extended Data Fig. 6f). Notably, one gene (CD276, encoding B7-H3) was significantly upregulated in TGFB2-rich regions.

Taken together, our results suggest that TGF-β2 is a major signalling ligand associating with impaired CD8+ T cell accumulation within SCC, acting in fibrovascular niches at the tumour-TME interface through epithelial-stromal-vascular cross-talk through the endoglin-ALK1 receptor complex. These areas are associated with local accumulation of high-risk histological features for metastasis and markers of immunotherapy resistance.

### Single-cell spatial profiling identifies a niche-specific role for TGF-β2 within the tumour microenvironment

To validate and expand further upon the role of TGF-β2 within immune-exclusive fibrovascular niches within CSCC, single-cell, image-based, spatial transcriptomic profiling of two CSCC was undertaken (Fig. 6a-b). Fluorescently barcoded RNA probes were identified and assigned to cells segmented by nuclear and membrane staining (Fig. 6c). A median (interquartile range) of 239 (137 - 405) transcripts across 112 (74 - 160) genes in 44707 cells were evaluated (Fig. 6d-g).

**Figure 6:**
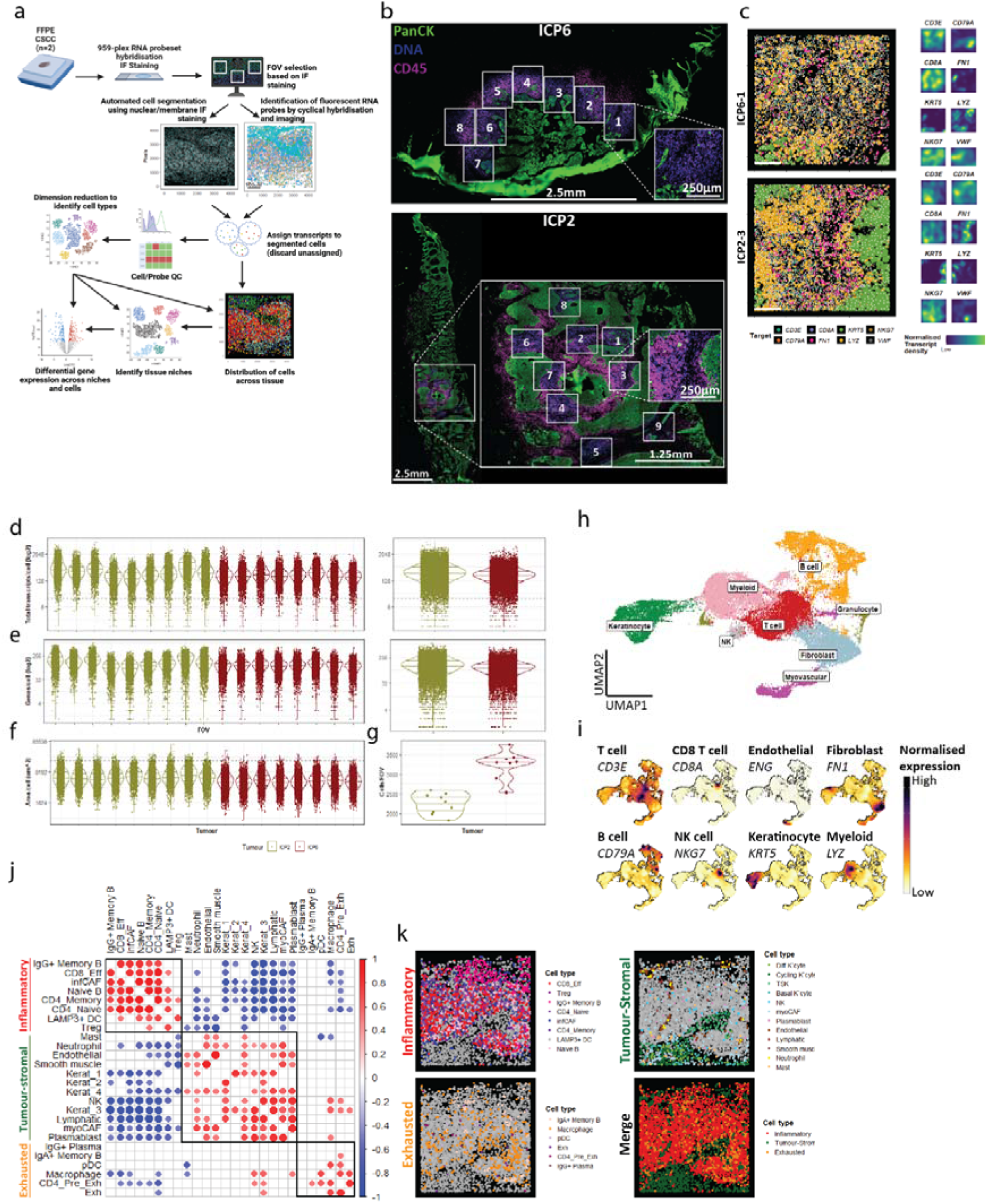
Workflow, CSCC and fields of view (FOV) used for single-cell spatial profiling. (A) Summary of workflow for acquisition and analysis; (B) Multiplex immunofluorescent images demonstrating FOV within CSCC (blue = nuclei, magenta = CD45, green = PanCK); (C) Left: Example of canonical cell marker transcript location within two FOV (white scale bar indicates 100µm); Right: normalised canonical cell marker transcript density within the same FOV (dark blue = low density, yellow = high density); (D) Total number of transcripts detected per cell stratified by FOV (left) and tumour (right); (E) Number of unique genes detected per cell, stratified by FOV (left) and tumour (right); (F) cell area, stratified by FOV. (G) Number of segmented cells detected per FOV, stratified by tumour. Black dashed line indicates QC threshold where applied. In plots D-G, yellow = FOV ICP2, red = ICP6. (H) UMAP dimension reduction plot illustrating broad cell type clusters based on transcriptomic profile (red = T cell, grey = NK cell, green = Keratinocyte, pink = myeloid, orange = B cell, magenta = myovascular, light blue = fibroblast. . (I) Smoothed canonical marker expression within UMAP space (white/yellow = low or no expression, black = high expression). (J) Spatial correlation of cell types within CSCC 50 nearest neighbour environments. Red = positive Spearman rank correlation within environments; blue negative Spearman rank correlation (black squares indicate hierarchical clusters forming three tissue compartments; orange = exhausted, red = inflammatory; green = tumour-stromal). (K) Spatial localisation of cell centroids within tissue compartments within a single FOV. Rowwise from left-to- right: inflammatory (shades of red/purple denote cell type within clusters identified in J – cells in other compartments denoted in grey); tumour-stromal (cells denoted by shades of green, blue, yellow and pink – cells within other compartments in grey); exhausted (cell types denoted by shades of orange, dark grey and brown – cells in other compartments in grey); Merged image denoting cells by compartment (red = inflammatory, orange = exhausted, green = tumour-stromal).

First-order cell populations within clusters were identified based on fluorescent staining and canonical gene expression (Fig. 6h-i and Extended Data Fig. 7a-b). The most common cell types identified were from the T cell, macrophage, stromal and epithelial populations (Extended Data Fig. 7c), with spatial distribution of these cells validating transcriptomic-derived identity (Extended Data Fig. 7d). Second-order clustering elucidated subsets based on canonical markers and module scores which were represented across all FOV from both tumours (Extended Data Fig. 8a-c). Two fibroblast populations were identified, each recapitulating transcriptomic features previously described in CSCC-associated inflammatory and myofibroblastic cancer-associated fibroblasts (infCAF and myoCAF respectively) [22] (Extended Data Fig. 8d). Four keratinocyte populations were annotated based on module scores of keratinocyte states, including one which enriched for genes consistent with the previously described ‘tumour-specific keratinocyte’ (TSK) [15] (Extended Data Fig. 8e). Nearest- neighbour tissue ‘environments’ clustered into three compartments: an inflammatory compartment, comprising naïve and effector T cells and some B cell subsets (Fig. 6j-k); a tumour-stromal compartment enriched for tumour, fibromyovascular, myeloid and granulocyte populations; and a more spatially disparate ‘exhausted’ compartment comprising exhausted and pre-exhausted T cells, macrophages and IgA+ memory B cells. Notably, CAFs demonstrated contrasting compartmental localisation, with infCAF associated with the inflammatory compartment and myoCAF enriched within the tumour-stromal compartment.

TGFB isoform transcripts exhibited distinct expression patterns between compartments; notably, whilst TGB1 and TGB3 expression was found across all three tissue compartments, TGFB2 expression was almost solely within tumour-stromal regions (Fig. 7a). TGFB2 was primarily expressed by tumour-specific keratinocytes (TSK) but was also present in other keratinocyte populations and myoCAF (Fig. 7b). TGF-β receptor expression exhibited similar heterogeneity: notably, endothelial cells were the sole expressors of ENG and expressed the highest levels of TGFBR2.

**Figure 7:**
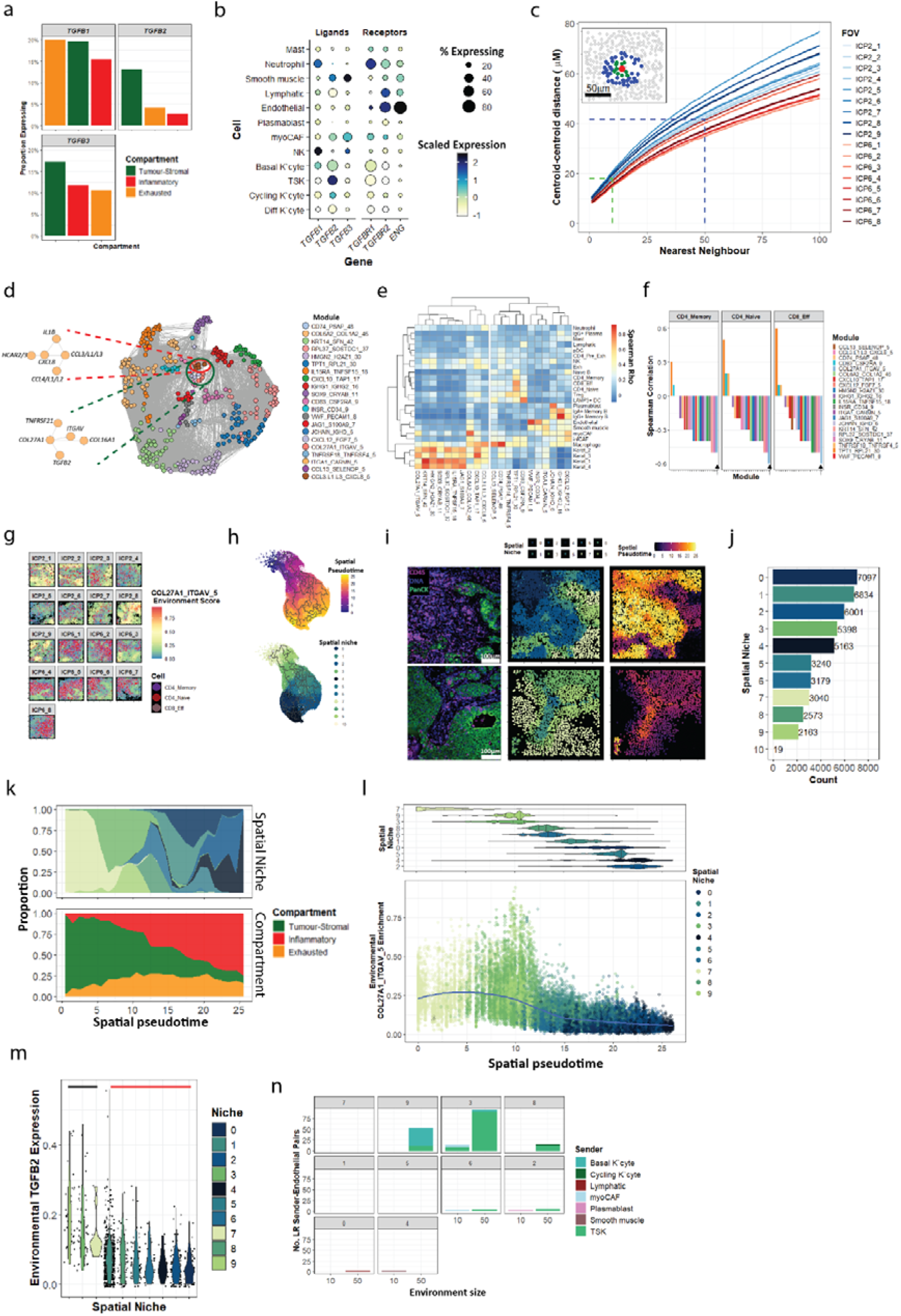
Spatial and transcriptomic profiling of TGFB signalling-related clusters. (A) Expression of TGFB ligand isoforms within tissue compartments (green = tumour-stromal, red = inflammatory; orange = exhausted). (B) Dot plot of TGFB isoforms and receptor expression within the tumour-stromal compartment. Dot size indicates proportion of the cell type with detectable expression, colour indicates scaled average expression (white = low expression, dark blue = high expression). (C) Centroid-centroid distance within each FOV for first 100 nearest neighbours. Dashed blue line indicates geometric mean distance to 50^th^ nearest neighbour, which was used to construct environments and for ligand-receptor interaction (LRI) analysis, whilst green dashed line indicates geometric mean distance to 10^th^ nearest neighbour, used in LRI analysis. Inset: example of 10 (green) and 50 (blue) nearest neighbours for red cell shown. (D) Adjacency graph delineating module construction from correlating genes. Inset: CCL3.L1.L3_CXCL8_5 module network (top, red lines); COL27A1_ITGAV_5 module network (bottom, green lines). (E) Heatmap of enrichment of co- expression modules within annotated cell populations (low correlation = blue, high correlation = red by Spearman rank test). (F) Spearman correlation of effector T cell populations (left panel CD4 memory, middle panel CD4 naïve, right panel CD8 Effector) with environmental module enrichment. The COL27A1_ITGAV_5 module is shown with a black arrow. (G) Spatial plot demonstrating enrichment of COL27A1_ITGAV_5 module (blue = low enrichment, red = high enrichment) and the relationship with effector infiltration (red spots = CD4 naïve, purple spots = CD4 memory, grey spots = CD8 effector) within FOV. (H) Spatial trajectory plot (origins indicated with white points, low pseudotime value = purple, high pseudotime value = yellow), bottom: spatial niches projected onto spatial trajectory plot. (I) IF image (Green = PanCK; Purple = CD45; Blue = DNA) (left panels) and spatial mapping of spatial niches (middle panels) and spatial pseudotime (right panels) at high power (FOVs shown ICP6-6 (top) and ICP2-8 (bottom)). (J) size of niches by cell count. (K) Evolution of spatial niche (top) and compartment (bottom) composition by pseudotime. Spatial niche colours are as in figure J. Compartment colours – red = inflammatory, green = tumour-stromal, orange = exhausted. (L) Top panel: violin plot of frequency of niche by pseudotime. Bottom panel: environmental enrichment of COL27A1_ITGAV_5 module with pseudotime. Blue line represents the fitted Locally Estimated Scatterplot Smoothing (LOESS) regression curve. (M) Environmental TGFB2 expression at 50nn resolution by niche (black and red lines indicate allocation of niches to ‘TGFB2-high’ and ‘TGFB2-low’ groups, respectively, for analysis in Fig. 8). (N) Frequency and sender cell of TGFB2 ligand-receptor interaction with endothelial cells by spatial niche at 10nn and 50nn resolution.

Genes were interrogated for spatial co-expression within 50-nearest neighbour (50nn) tissue ‘environments’ (Fig. 7c). Genes exhibiting cell-independent spatial co-regulation (n=364) were constructed into 21 clusters (Fig. 7d and Extended Data Fig. 9a). TGFB2 formed a spatial co-regulation cluster with four other genes associated with extracellular matrix turnover (ITGAV, COL27A1, COL16A1 and TNFRSF21), which were enriched within and around keratinocytes and myoCAF (Fig. 7d-e and Extended Data Fig. 9b). Notably, this module (COL27A1_ITGAV_5) demonstrated the strongest inverse relationship with effector T cell proportion within environments, which was evident spatially across both tumours (Fig. 7f-g and Extended Data Fig 9d).

Clustering of 50nn environments by cell population permitted identification of ten CSCC ‘niches’ conserved across both tumours (Fig. 7h and Extended Data 9e). Unidimensional spatial trajectory analysis of cell environments revealed a progression of cell types from within the keratinocyte- dominated tumour bed with little effector T cell infiltration (niche 7), through a transitional zone representing the leading tumour edge (niches 3 and 9), into the remaining immune-dominated niches within the surrounding stroma (Fig. 7i-k and Extended Data Fig. 9f-g). A combination of spatial niche and pseudotime were used to represent distinct tumour subregions and a continuous, unidimensional representation of distance to the local tumour bed, respectively. Whilst most cell populations demonstrated niche-specific enrichment, it is notable that macrophages were uniformly present across all tissue regions (Extended Data Fig. 9g).

Greatest COL27A1_ITGAV_5 environmental enrichment was seen in the pseudotime range equating to the transition zone (Fig. 7l), equating to niches 3 and 9 (Fig. 7m). Accordingly. evaluation of TGFB2- related cell-cell and environmental signalling (at 10 and 50nn resolution, respectively) revealed TGFB2 LRI with endothelial cells were concentrated in niche 3 and niche 9, predominantly arising from TSK and myoCAF (Fig. 7n). This was consistent with the earlier histological findings that TGF-β2 staining is concentrated at the leading tumour-edge.

### Inflammatory signalling primes TGF-β2-driven endothelial-mesenchymal transition at the leading tumour edge

We next focused upon the downstream effect of TGF-β2 signalling upon the endothelium. Endothelial cells within TGFB2-rich niches at the leading tumour edge (Fig. 7m) exhibited upregulation of genes and gene sets relating to ECM production and turnover, including collagens and fibroblast markers such as S100A4 (Fibroblast-specific protein, FSP) compared to endothelial cells in tumour-distant niches (Fig. 8a-b). Notably, upregulated transcripts included ACVRL1 (encoding ALK1), IL32 (IL-32), LGALS1 (Galectin-1), CYTOR (Linc00152) and ETS1, which have been associated with epithelial-mesenchymal transition, tumour progression and poor prognosis in squamous cell carcinoma at other sites [23–28]. 16 of 23 differentially expressed genes were core genes within the Angiogenesis/ECM module previously identified by bulk spatial transcriptomic profiling, with LGALS1 having the strongest module membership (Supplemental Data). Endothelial cells within TGFB2-rich niches demonstrated upregulation of myoCAF but not iCAF gene module expression (Extended Data Fig. 10a). Within endothelial cells, myoCAF gene module enrichment demonstrated positive correlation with the COL6A2_COL1A2_46 spatial module and inverse correlation with the vasculature gene-related VWF_PECAM1_8 module (Extended Data Fig. 10a). Taken together, this indicates TGFB2 expression at the leading tumour edge is associated with local trans-differentiation of endothelial cells into myoCAF.

**Figure 8:**
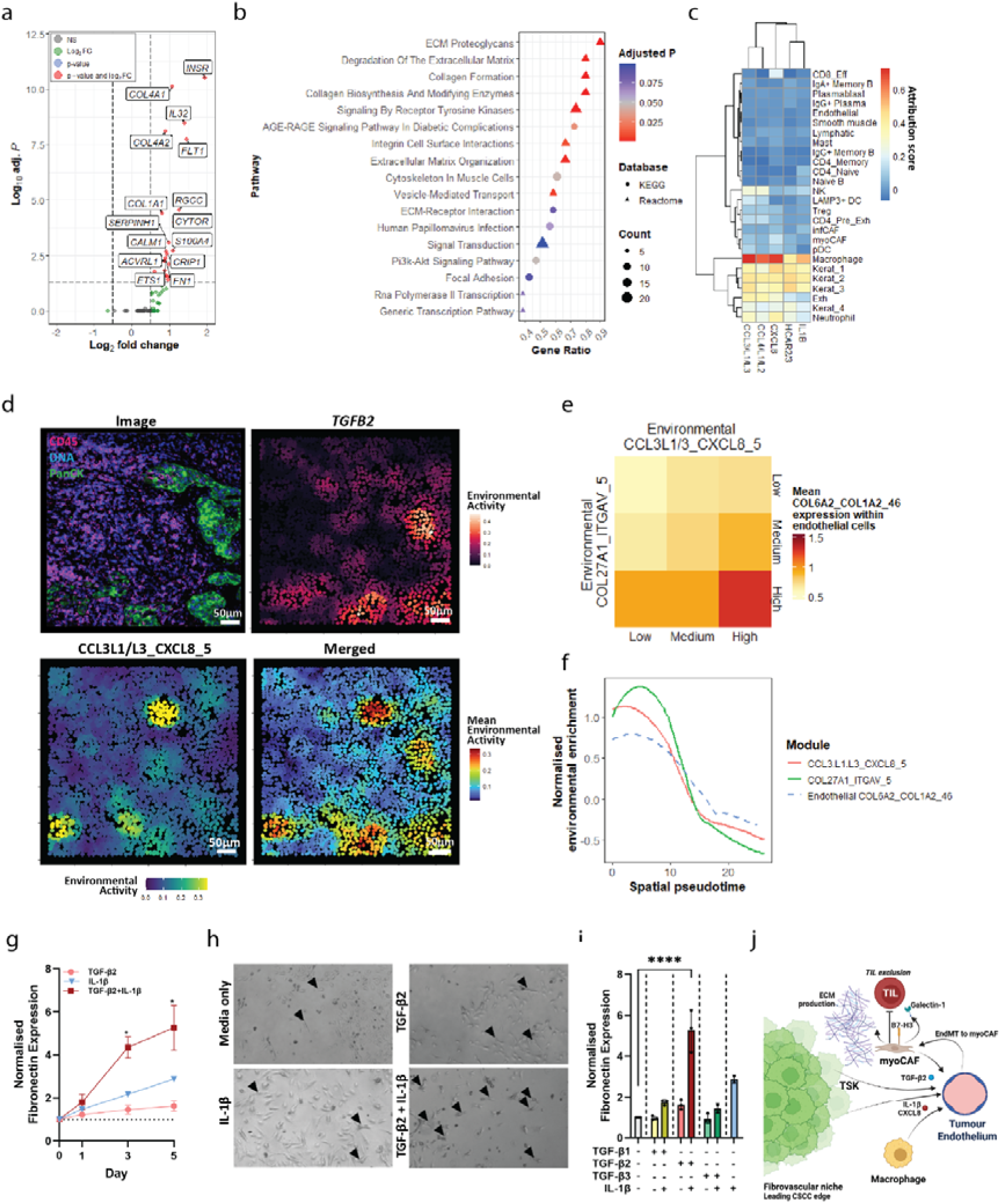
Effect of TGF-β2 upon endothelial cells (A) Volcano plot of differentially expressed genes in endothelial cells from TGFB2-high and low niches. Absolute LogFC and adjusted p value thresholds were set at 0.5 and 0.05 respectively. Red spots = genes with log-fold change >0.5 and adjusted p value <0.05; green spots = log fold change >0.5 and adjusted p value >0.05; grey spots log fold change <0.5 and adjusted p value >0.05. (B) Gene sets significantly enriched within endothelial cells from TGFB2-rich niches compared to those from TGFB2-sparse niches. Y axis indicates gene ratio; spot size gene count; colour adjusted p value (blue indicates adjusted p value >0.05, red adjusted p value <0.05). (C) Attribution score (Spearman rank correlation) of genes within CXCL8_CCL3.L1.L3_46 module by cell type (blue = low attribution; red = high attribution). (D) Expression of TGFB2 and IL1B in a single typical FOV (row-wise from top-left, IF image; environmental TGFB2 expression (low expression dark red, high expression light red); environmental IL1B expression (low expression dark blue, high expression yellow); summed IL1B+TGFB2 expression (low expression dark blue, high expression dark red) (E) Mean expression of COL6A2_COL1A2_46 module within endothelial cells, stratified by environmental expression of COL27A1_ITGAV_5 and CCL3L1/3_CXCL8_5 gene modules (divided into tertiles of expression – low COL6A2_COL1A2_46 expression = yellow, high expression = orange-red). (F) Environmental expression of COL27A1_ITGAV_5 (green line) and CCL3.L1.L3_CXCL8_5 modules in spatial pseudotime. Dashed blue line indicates expression of the COL6A2_COL1A2_46 gene module within endothelial cells. (G) Time course of fibronectin expression in human umbilical vein endothelial cells (HUVEC) in culture with 10ng/mL of specified cytokines and detected by flow cytometry. Pink line TGF-β2 only; blue line IL-1β only; red both TGF-β2 and IL-1β * p<0.05 compared to baseline, by mixed effect modelling with Geisser-Greenhouse correction, for culture condition and timepoint, blocking by experimental replicate, following Dunnett’s multiple comparison test. Dotted line indicates unstimulated expression. Results represent at least three independent experiments. (H) High power microscopy of HUVEC after five days in culture with (clockwise from top left) media only, 10ng/mL TGF-β2, 10ng/mL TGF-β2 and IL-1β, 10ng/mL IL-1β. Black triangle indicates HUVEC developing an elongated spindle morphology. (I) Expression of fibronectin following 5 days of culture with 10ng/mL of specified cytokine(s) and detected by flow cytometry. Cytokine presence within culture condition indicated with ‘+’. **** p<0.0001 by by ANOVA with post-hoc Šídák’s multiple comparisons test against control (media only). (J) Conceptual model of local signalling networks driving endothelial trans-differentiation to myoCAF within fibrovascular niches.

Notably, the COL27A1_ITGAV_5 module was closely spatially co-regulated with a second module, CCL3.L1.L3_CXCL8_5 (Fig. 7d), consisting of genes relating to innate inflammatory signalling and produced predominantly by macrophages and the tumour bed (Fig. 7e and 8c). This was validated by niche-stratified ligand-receptor interaction signalling, demonstrating most LRI arising from these cells and this module in niches 3 and 9 (Fig. 8d and Extended Data Fig. 10c). We hypothesised that local inflammatory signalling may contribute, though not drive, this process, as evidenced by a synergistic effect of environmental expression of both modules upon endothelial expression of the myoCAF- associated COL6A2_COL1A2_46 module, particularly at the leading tumour edge (Fig. 8d-f).

Functional validation of the synergistic effect of TGF-β2 and local inflammation upon endothelial cell behaviour was undertaken using Human Umbilical Vein Endothelial Cells (HUVEC). In the presence of both TGF-β2 and IL-1β, HUVEC markedly upregulated fibronectin expression (Fig. 8g) and developed altered cell morphology, adopting a spindle-like appearance (Fig. 8h). This effect was accompanied by upregulation of other fibroblast markers and was isoform specific, as this was not seen with TGF-β1 or TGF-β3 (Fig. 8i and Extended Data Fig. 10d). Upregulation of CD276 (B7-H3) was driven by IL-1β in a dose-dependent manner (Extended Data Fig. 10e).

## Discussion

CSCC represents a major and increasing cause of morbidity and mortality worldwide. CSCC demonstrates a range of histopathology, with significant intratumoral heterogeneity considered a high-risk feature associated with poorer outcome [29]. The incomplete success of current immunotherapy regimens creates a need to identify pathways underlying tumour escape from TIL- driven responses.

To address this, we utilised multimodal spatial profiling to evaluate signalling networks orchestrating TIL behaviour within the CSCC TME. We identify a major role for the tumour-stromal compartment in creating TME niches which exclude TIL and may associate with poorer response to immunotherapy.

Single-cell spatial profiling and functional validation identified TGF-β2 as a hub soluble factor orchestrating the heterogeneity of immune response by priming endothelial cells to transdifferentiate to suppressive cancer-associated myofibroblasts in an inflammatory setting at the leading tumour edge.

Whilst it has been long known that the TME around TILs contains a multitude of stromal populations [30], the pathways by which these cells acquire suppressive characteristics have been less explored. Accumulation of CAF due to epidermal-mesenchymal transformation (EMT) is well-described [31], and has been specifically attributed to the tumour-specific keratinocyte (TSK) subpopulation in CSCC [32]. Our results suggest that TSK may also orchestrate mesenchymal transition in local vascular populations, which may be an underappreciated and a stronger driver of TIL behaviour than previously acknowledged. Technical and amplification bias means spatially agnostic single-cell RNA sequencing (scRNAseq) approaches often enrich against stromal populations: for example, less than 10% of the scRNAseq dataset used by Ji et al comprised endothelial cells and fibroblasts [15]. Our data suggest that areas rich in fibrovascular cells and transcriptional activity were associated with the lowest infiltration of TIL. Ji et al used multimodal profiling to identify fibrovascular niches within CSCC, predominantly consisting of TSK, CAF and endothelial cells [15]. TSK were postulated to represent hub signalling nodes within such niches, but the study was limited in its ability to identify drivers of such behaviour. Our findings build upon and expand our understanding of fibrovascular niche biology. Targeting of fibrovascular signalling may disrupt niche development, enriching local immune responses and slowing tumour invasion and metastasis.

Here, cross-correlated multimodal analysis now shows TGF-β2, predominantly secreted by TSK and fibroblasts, as the ligand with the strongest negative association with CD8+ TIL accumulation. The pleiomorphic role of TGF-β signalling during CSCC progression has been well studied, particularly with respect to TGF-β1. These have revealed a progressive switch from a tumour suppressor to a tumour promoter role with establishment of the CSCC microenvironment [15, 33, 34]. Our data reveal a hitherto unappreciated niche-specific role for TGF-β2 in driving endothelial transdifferentiation, CAF expansion and controlling CD8+ TIL accumulation at the leading tumour edge.

Interestingly, on transcriptomic LRI evaluation TGF-β2 was not found to associate with betaglycan (TGFBR3), the co-receptor with highest affinity for this TGF-β isoform [35], instead associating with endoglin (CD105), which classically exhibits low binding affinity for TGF-β2 [36]. Two possibilities may explain this. Firstly, TGFBR3 expression correlates poorly with protein levels, potentially obscuring an interaction [37]. Secondly, betaglycan may form complexes with endoglin on the surface of endothelial cell lines in a ligand-dependent and -independent manner prior to recruitment of TGFBRII [38]. Endoglin may therefore not directly bind TGF-β2 but instead form part of the signalling complex leading to modulation of downstream signalling and endothelial-mesenchymal transition (EndMT) induction.

Spatial co-expression of TGFB2 with a module of genes including ITGAV, COL27A1 and COL16A1 was seen, with strongest environmental expression around the leading tumour edge. Expression of ITGAV, encoding integrin α_V_, has been associated with immunotherapy resistance in lung cancer through activation of TGF-β [39]. Pro-TGF-β2 is activated by integrin α_V_β_6_ in vitro [40], indicating this module may drive local activation of ECM-bound latent TGF-β2. COL16A1 and COL27A1, encoding collagen types XVI and XXVII alpha 1 chains respectively, have been identified as expressed by the stroma in other malignancy and in cardiac fibrotic remodelling [41, 42]. Whilst the significance of COL27A1 expression is unclear, expression of COL16A1, a linker molecule which regulates stability within the ECM, is a poor prognostic feature in oral SCC, leading to activation of MMP9 and increased tumour invasiveness [43].

ITGAV_COL27A1_5 module expression aligned spatially with a module constituting innate inflammatory signals, with environmental co-expression of both modules associated with endothelial upregulation of a myoCAF-like gene module and pathways relating to ECM turnover. Functional validation confirmed that TGF-β2 was able to drive this effect, but only in the presence of inflammatory signalling via IL-1β. CXCL8 similarly drives EndMT through NFκB signalling pathways, likely representing a common final pathway through which innate-signalling cytokines exert this effect [44]. Animal models using lineage reporters have suggesting up to 40% of CAF may derive from the endothelial pool [45]. Here, for the first time, we provide detailed in vivo evidence for the importance of this process in one of the most common cancers in the developed world, and tie this directly to local suppression of TIL accumulation.

MyoCAF and transdifferentiating endothelial cells demonstrated upregulation of LGALS1 expression, encoding for Galectin-1, which was also a hub gene within the Angiogenesis/ECM remodelling module and was strongly negatively associated with CD8+ T cell infiltration on bulk spatial transcriptomic analysis. Galectin-1 is a β-galactoside-binding protein with multiple effects within the TME, including promotion of angiogenesis and immunoregulation and apoptosis in effector T cells, leading to immune escape [46]. Fibrovascular niches were associated with reduced expression of various components of the tumour inflammation score (TIS), which has been validated as predicting response to checkpoint blockade therapy across multiple malignancies, although not CSCC [21]. Increased expression of markers consistent with TSK have been demonstrated to predict poorer response to immunotherapy in CSCC [15]. Whilst our study evaluated only primary lesions and not metastases, we hypothesise that the TIS could predict response to immunotherapy in patients with advanced CSCC also. Fibrovascular niches express little PD1 or CTLA4, but did express high levels of CD276 (coding for B7-H3), which was confirmed in vitro to be driven by inflammatory signalling, and correlated negatively with CD8 accumulation. The natural receptor for B7-H3 is unknown [47], meaning full ligand-receptor interaction analysis could not be performed in our study. However, expression in cancer tissue is associated with suppressed CD8+ responses and poorer disease outcomes [48], and may provide another mechanism by which myoCAF suppress local TIL accumulation [49]. B7-H3 also has a role in driving tumour proliferation and metastasis, with clinical trials underway regarding the therapeutic potential of blocking this pathway [47]. This indicates fibrovascular niches may be responsive to alternative or combined checkpoint blockade therapy.

Our findings build on those previously reported, allowing construction of a conceptual model (summarised in Fig. 8j). Tumour-stromal derived TGF-β2, activated by local integrin α_V_, primes local endothelial cells to transdifferentiate into mesenchymal populations demonstrating transcriptomic hallmarks of suppressive myoCAF in the presence of inflammatory signalling from infiltrating macrophage populations, which inhibit TIL accumulation in a Galectin-1 and B7-H3 dependent manner. This is particularly pronounced at the leading tumour edge as this represents the interface between tumour-driven signalling and inflammatory signalling from local leucocyte populations. TGF- β2 staining suggests this process may become more pronounced as the tumour evolves.

It is important to be aware of limitations in this study. The clinical effects of our findings remain to be tested, though TGF-β2 expression correlated positively with increasing histological stage, which is associated with poorer outcomes [16]. Evaluation through publicly available datasets such as The Cancer Genome Atlas are hampered by the lack of a CSCC dataset and bulk transcriptomic data, which may miss niche-specific effects. Whilst our sample size submitted for spatial profiling is relatively small, external validation by immunoenzymatic profiling and by using publicly available CSCC datasets increases the confidence of our findings.

In summary, we have used multimodal spatial profiling with functional validation to delineate a major signalling network driving the establishment of immune-sparse fibrovascular niches at the leading edge of CSCC, potentially clearing a path to facilitate tumour cell invasion. This identifies a central role for synergistic inflammatory-TGF-β2 signalling leading to endothelial-mesenchymal differentiation of the local vasculature and accumulation of inhibitory myoCAF. Such niches demonstrate multiple markers of angiogenesis, metastasis and checkpoint therapy resistance. Future work should focus on methods to disrupt these signalling networks.

## Supporting information

Differentially Expressed Genes in Immune vs Tumour Rich Regions

Gene WCGNA Module Membership and Association with Histology

Pathway Enrichment Within WCGNA Modules

Ligand-Receptor Interactions within Immune-Rich Regions

InSituCor Module Genes

WGCNA Genes Upregulated in Endothelial Cells in TGFB2-High Niches

## Data Availability

All data produced in the present study are available upon reasonable request to the authors

## Acknowledgements

The authors would like to thank the staff and patients at the Oxford Dermatology and Transplant Units for their support of this study, in particular R. Matin for her assistance in recruitment of non-immunosuppressed participants. We are grateful to M. Veretennikova (CAMS Oxford Institute) for her bioinformatic support. This work was supported by grants from the British Skin Foundation, Oxford University Hospitals Charitable Fund, Wellcome Trust (grant reference: 098744/Z/12/Z) and the Chinese Academy of Medical Sciences (CAMS) Innovation Fund for Medical Science (CIFMS), China (grant number: 2018-I2M-2-002), Oxford NIHR Biomedical Research Centre and Medical Research Council UK.

The authors acknowledge the support of the National Institute for Health Research, through the Local Clinical Research Network. Figures delineating experimental workflow and the conceptual model were generated using Biorender. MM, TMV and LLB are employees of Bruker Spatial Biology, the manufacturers of the GeoMx and CosMx platforms used in this study. The remaining authors declare no relevant conflicts of interest.

## Methods

### Participant recruitment and sampling

Participants with a recent history of CSCC but no history of continuous systemic immunosuppression (immunocompetent, ICP), were recruited from the Outpatient Department at the Oxford Dermatology Unit, Churchill Hospital. Kidney transplant recipients (KTR) with stable graft function and a history of CSCC were recruited during routine transplant clinic follow-up at the Oxford Transplant Unit. Potential participants were ineligible if they had evidence of previous chronic hepatitis or HIV infection, or a history of non-keratinocyte malignancy within the last five years. Recruitment was part of a longitudinal study of immunological and clinical risk factors for malignancy development in long-term KTR [50, 51].

This conduct of this study was reviewed and approved by the West of Scotland NHS Research Ethics Committee (Reference 12/WS/0288) and the study was undertaken in accordance with the principles of the Declaration of Helsinki. Participants provided written, informed consent prior to participation.

### Histological and immunoenzymatic profiling

Intact formalin-fixed, paraffin-embedded (FFPE) CSCC from study participants which were previously excised as part of routine clinical practice were utilised.

Archived samples were re-reviewed by a dermatopathologist to confirm the clinical diagnosis of CSCC and histological features described in Table 1. Intratumoral and peritumoral CD8+ density were evaluated by immunohistochemistry. 3-4µm sections were cut on coated SuperFrost Plus slides (Thermo Fisher Scientific), prior to staining.

### CD8 staining

Double staining utilised 3,3’-diaminobenzidine (DAB, brown) for CD57 (NK and NKT cells) and alkaline phosphatase (red) for CD8 staining. Epitope retrieval for CD57/CD8 was performed with Bond Epitope Retrieval Solution 1 (Citrate-based buffer at pH 6.0) at 100 c for 20 minutes. CD8 detection was performed using ‘Bond Polymer Refine Red Detection System’ (Leica Biosystems). Sections were counterstained with haematoxylin then mounted with aqueous mountant.

The sections were stained on the Leica Bond Max staining platform (Leica Biosystems, UK). The CD8 and CD57 primary antibodies (clones 4B11 and NK-1, Leica Biosystems, respectively) were used as supplied and in accordance with manufacturer’s recommendations.

Cells were counted and summated over 10 high power fields (HPF) by a dermatopathologist (IE), equating to a total surface area of 2mm^2^ . These were then adjusted to give a value per mm .

### CD8/TGF- β2 co-staining

Manual double staining (Abcam, ab210059) for TGF-β2 and CD8 on FFPE CSCC sections (n=27) was performed according to manufacturer’s protocol. Briefly, after deparaffinisation and rehydration slides were permeabilised prior to antigen retrieval using IHC Antigen Retrieval Solution (10x High pH, Invitrogen, 00-4956-58). Endogenous peroxidase activity was blocked using hydrogen peroxide prior to incubation with polyclonal mouse TGF-β2 (Abcam, Ab167655) and rabbit CD8 (Abcam, AB4055) primary antibodies. Following washing, slides were incubated with rabbit alkaline phosphatase (AP) Polymer and mouse horseradish peroxidase (HRP) Polymer prior to colour development using DAB and Permanent Red Working Solutions, with nuclear counterstaining using Mayer’s Hematoxylin Solution (Sigma Aldrich).

Slides were visualised using EVOS M5000 widefield microscope, with staining quantified using FIJI (ImageJ). CD8+ and TGFB2+ cells were manually counted over 5x 200X images of the leading tumour edge for each CSCC.

### Immunofluorescent and GeoMx Digital Spatial Profiling

Slides were processed in accordance with manufacturer’s instructions. Freshly sectioned paraffin- embedded samples, some dual loaded onto slides (Fig. 1B) were submitted to deparaffinization, rehydration, heat-induced epitope retrieval (for 20 minutes at 100°C), and enzymatic digestion (1 μg/mL proteinase K for 15 minutes at 37°C), performed on the Leica BOND-RX. Tissues were incubated with 10% neutral buffered formalin for 5 minutes followed by 5 minutes with NBF Stop buffer. The tissue sections were hybridized with the oligonucleotide probe mix (Whole Transcriptome Atlas) overnight, then blocked and incubated with PanCK-AF532 (clone AE1+AE3; Novus), CD8-AF647 (clone OTI3H6; Origene), CD57-AF594 (clone HNK-1; Biolegend), and DNA dye (Syto13-AF488 dye; Invitrogen) for 1 hour. Tissue sections were loaded into the Bruker Spatial Biology (BSB) GeoMx™ DSP platform and high-resolution images acquired using proprietary software, prior to region selection for transcriptomic analysis [52]. Polygonal areas of interest (AOI) were chosen based on morphology and staining pattern, with peritumoral regions rich in CD8 staining taken to represent immune-rich regions (IRR) and regions rich in panCK staining with irregular and malignant-appearing morphology taken to represent tumour-rich regions (TRR). Where feasible, IRR and TRR were chosen based on histological proximity. Polygons were generated to avoid contamination by other cell types (for example, avoiding PanCK-positive nests in IRR) whilst maximising the number of desired cells (for example, CD8-positive cells in IRR) within the polygon boundary (limited to 680x785µm).

UV light directed at each AOI in turn released oligonucleotides that were collected and prepared for sequencing. Illumina i5 and i7 dual-indexing primers were added during PCR (4 μL of collected oligonucleotide per AOI) to uniquely index each AOI. The PCR parameters were specified by BSB (NGS Readout Reference Manual MAN-10153). AMPure XP beads (Beckman Coulter) were used for amplicon purification. Library concentration as measured using a Qubit fluorometer (Thermo Fisher Scientific), and quality was assessed by High Sensitivity DNA ScreenTape Analysis (Agilent). Sequencing was performed on an Illumina Novaseq 6000, undertaken in-house at BSB, and FASTQ files were processed by the DND pipeline, resulting in count data for each target probe in each AOI. A no-template control sample was sent for sequencing to exclude exogenous contamination.

### Histological evaluation of AOI

Histological and immunofluorescent characteristics of AOI submitted for transcriptomic evaluation, including number of nuclei and absolute number of nucleated CD8+ and panCK+ cells, were derived using a custom pipeline on CellProfiler software [53]. Intensity thresholds were set for object identification of nuclei and panCK+ cells, with CD8+ cells identified by adapted intensity thresholds. Visual validation of object identification for each AOI was performed to confirm accuracy of identified objects. AOI size, quantified by GeoMx software at time of AOI selection, was used to generate panCK+ and CD8+ cell density from cell number within each AOI. CD8+ and panCK+ cell proportion was calculated as the number of CD8 or panCK+ cell objects divided by the total number of nuclei identified within that AOI. Nuclei count and surface area obtained via Cellprofiler analysis demonstrated strong correlation with those enumerated on the GeoMx DSP (Fig. S3A-C).

### GeoMx Analysis

### Quality Control and Normalisation

Quality control and normalisation were undertaken using the GeoMxTools v1.5.0 R package. QC thresholds at an AOI level were:

- Over 80% of reads trimmed, stitched and aligned per AOI
- >1000 reads per AOI
- Proportion of unique probe reads of >50% of total reads per AOI.
- >100 nuclei per AOI

All AOI achieved these thresholds. A pool of negative probes was used to quantify background signal, with the geometric mean of negative probe counts multiplied by the square of the geometric standard deviation set as the ‘limit of quantification (LOQ)’. Probes representing genes detected above the LOQ in less than 5% of AOI were excluded from further analysis (n=5,743 genes). After probe-level quality control, a total of 12,934 genes were used for downstream evaluation (Fig. S4A), with an n+1 adjustment for any genes with zero expression counts within AOI to facilitate downstream transformation. Normalisation to account for inter-region differences in cellularity and transcriptomic activity was undertaken using the upper-quartile (Q3) method. Normalisation using this approach demonstrated good segregation and linearity of Q3-normalised and negative control counts (Fig. S4B-C). Further evaluation of the adequacy of normalisation was undertaken using negative control probe counts and a panel of 37 housekeeping genes (Fig S4D-E). The genes used for this evaluation were TUBB, RAB7A, HDAC3, DNAJC14, POLR1B, TBC1D10B, TBP, TMUB2, NRDE2, TLK2, G6PD, MRPL19, STK11IP, PUM1, SDHA, ABCF1, SF3A1, GPI, PSMC4, POLR2A, TPM4, UBB, RPS29, PSMB2, EIF2B4, ERCC3, TFRC, GUSB, RPLP0, OAZ1, HPRT1, RPLP0, TBPL2, ARPC2, ZNF410, PUM1 and PPIA.

### Dimension Reduction

Dimension reduction was undertaken using principal component (PC) analysis of normalised data; the first four dimensions were chosen for evaluation based on the elbow method (Fig. S5A), which cumulatively account for 38.3% of transcriptomic variability across AOI. Other potential sources of variation within these first four PC were evaluated (Fig. S5B-G). Whilst slide represented an additional source of transcriptomic variation, ‘Tumour’ and ‘Slide’ variables demonstrated significant collinearity (Fig. S5J) and so ‘Donor’ was chosen for linear modelling, representing the more biologically relevant variable.

### Differential gene expression

Differential gene expression between IRR and TRR was performed using the ‘differential expression for repeated measures (dream)’ approach (variancePartition and EdgeR packages) [54], modelling for AOI type (fixed effect) and donor (random effect). Significance was taken as an Benjamini-Hochberg adjusted P value of <0.05 and an absolute log_2_ fold change of >0.6 (i.e. 1.5-fold absolute change).

### Gene Set and Pathway Analysis

Gene Set Enrichment Analysis was performed on all genes ranked by log-fold change between IRR/TRR using the Gene Ontology ‘biological processes’ knowledgebase, with a minimum/maximum gene set size of 30 and 500 genes respectively (clusterProfiler package). Redundant pathways, defined as those with a semantic similarity above 0.7, were removed using the simplify function.

Pathway analysis was undertaken using Log_2_ transformed counts and the ‘Pathway Analysis with Downweighting of Overlapping Genes’ (PADOG) method, which reduces the influence of genes common to multiple pathways [17], and the ReactomeGSA package. The Reactome knowledgebase was interrogated, with exclusion of disease pathways and any pathways with less than 30 genes.

### Cellular Deconvolution

Spatial deconvolution was undertaken using the SpatialDecon R package as detailed by Danaher et al and previously validated on data generated from the GeoMx platform [18]. Briefly, log-normal regression using a reference cell matrix was used to infer abundance of previously described cell populations within AOI. The reference cell matrix used for deconvolution was derived from publicly available data from cell populations identified by single-cell transcriptomic analysis of human CSCC [15] and available at Gene Expression Omnibus (GEO) reference GSE144240. Raw and Q3 normalised counts, background counts (from negative control probes) and the CSCC reference matrix were used as input. No adjustment for inferred tumour profile was made, as TRR and IRR were treated as separate entities and panCK-staining cells were deliberately excluded from IRR; supporting this approach, adjustment for tumour-profile did not change estimates of cell abundance (data not shown). Cell types with a summed abundance score of zero across all ROI were excluded from downstream analysis.

### Weighted Gene Co-expression Network Analysis (WCGNA)

WGCNA (WGCNA package) was undertaken using normalised genes detected above the LOQ. Only IRR were used, given the significant transcriptional heterogeneity between IRR and TRR. Hierarchical clustering by average linkage did not reveal any outliers so all IRR were included. The soft- thresholding power was 5, based upon plateau of the scale-free topology fit index curve at >0.85 and mean connectivity of less than 50 (Fig. S7a-b). ‘Bicor’ method of signed hybrid correlation was used, with a limit upon excluded outliers of 10%. A signed topological overlay matrix type was constructed, with hierarchical clustering on dissimilarity in this matrix by average linkage method to generate modules. The minimum module size was 30 genes, with a dendrogram cut height of 0.3 (corresponding to a correlation of 0.7) for merging of similar modules (based on dissimilarity of module eigengenes). A single gene (TM6SF1) was unassigned following this process and was discarded for downstream analysis.

### Gene network annotation

Modules identified through WGCNA were correlated (using Spearman rank test) with inferred cell populations, generated by spatial deconvolution. Functional enrichment analysis was performed for each module’s gene list, interrogating the Reactome, KEGG and Gene Ontology (Biological Processes subontology) knowledgebases, using g:Profiler (version e110_eg57_p18_4b54a898 and the gprofiler R interface) with FDR multiple testing correction method applying a significance threshold of 0.01 [55]. Enriched pathways and correlated cell populations were used to manually annotate gene networks by biological theme. Modules with no enriched gene sets were excluded from annotation.

Module membership was calculated by correlation of the module eigengene and gene expression values across IRR ROI. Gene significance was calculated as the absolute correlation between CD8 density or proportion of nucleated cells and gene expression across IRR ROI.

### Ligand-receptor interaction profiling

Ligand (L)-receptor (R) interaction (LRI) profiling within IRR was undertaken using the BulkSignalR approach [56]. The issue of background noise complicating LRI deconvolution in bulk data is accounted for by incorporating not only LRI but also downstream pathway (pw) alterations in gene targets (T). A null distribution of L – R Spearman rank correlations is generated for all possible upper-quartile normalised L-R combinations. L-R correlation significance is obtained directly from the null distribution. However, a further step is performed by correlating R with all downstream T with significance evaluated by order statistics. Thus L, R and pw must demonstrate correlated activities to be deemed significant.

LRI were taken from the LRdb knowledgebase, whilst pathway information was obtained from the Reactome and Gene Ontology databases. The statistical training model parameters were adjusted to account for shallower data with reduced dynamic range present within spatial transcriptomic profiling datasets: namely a minimum correlation threshold of -1, at least 2 targets per pathway within the dataset, and a Q-value threshold of 0.05 [56]. Promiscuity of ligands for multiple receptors was addressed by reduction to the top-correlated interacting receptor (based on smallest p value calculated by Spearman rank test); a similar approach was used to reduce redundancy and collapse multiple downstream pathways. To focus analysis upon the most relevant pathways within TME, only L-R with at least one moderate and significant positive interaction (defined as an LR correlation of >0.3 and triple q-value of <0.05) were included for final analysis.

### Integration of histological data

CD8+ T cell density and proportion for each IRR was correlated to transcriptomic features initially by Spearman’s’ rank correlation and then subsequently by mixed effect modelling, treating participant ID as a random effect. P values were adjusted for multiple testing as previously described.

### Single-cell transcriptomic profiling

### Sample preparation and data acquisition

Tumours for single-cell profiling were chosen based on being representative of the samples evaluated via GeoMx, and based on significant heterogeneity of CD8+ accumulation as evaluated in Fig. 1G-H.

Samples were prepared for analysis on the CosMx™ SMI platform according to manufacturer’s instructions (detailed in MAN-10159-01, available from BSB). Briefly, freshly sectioned 5µm sections of two CSCC (ICP2 and ICP6) were deparaffinised and rehydrated prior to heat-induced epitope retrieval, enzymatic digestion (3 μg/mL proteinase K for 30 minutes at 37°C) and fiducial application. The Human Universal Cell Characterisation Panel (BSB, Seattle, US) probeset was hybridised overnight before stringent washing and staining for nuclear (DAPI) and membrane (CD298/β2M) segmentation markers, and CD45 and PanCK (CosMx™ Human Universal Cell Segmentation and IO PanCK/CD45 Supplemental Segmentation Kits respectively, BSB).

Prepared slides were run immediately on the CosMx™ SMI Platform, which utilises automated cycled in situ hybridisation, imaging and cleavage of reporter probes [57]. Nuclear and membrane staining is used to create cell boundaries using a modified Cellpose neural network approach, with fluorescent transcripts identified as falling within those bounds assigned to each cell (segmentation). Transcripts not assigned to a cell are excluded from further analysis. Regions for analysis (fields of view, FOV) were chosen to represent regions of tumour microenvironment; ICP6 was sufficiently small that all TME was acquired, whilst ICP2 was a larger tumour and so representative areas of tumour/stroma were chosen, based on PanCK/CD45 staining.

### Quality control

Other QC parameters applied were used to exclude cells with: <20 detected transcripts; >10% of detected transcripts were negative control probes; the ratio of total transcripts to unique transcripts <1.2; calculated cell area > 75 centile plus 3 x interquartile range across all cells (i.e. the 3IQR method, equating to a cut-off of 17454um^2^). 1535 cells (3.3% of pre-QC total) were excluded. Field of view (FOV) QC, performed by evaluation of FOV sub-fields (49 per FOV) for overall loss of signal, or loss within any barcode (failed reporter cycles), did not flag any FOV or genes for exclusion.

### Integration, normalisation and sequential dimension reduction for population identification

The dataset from each tumour was merged using the Seurat v5 workflow for integration, and downstream processing and dimension reduction took place within the Seurat environment. Briefly, each tumour was normalised individually with variance stabilisation using Pearson residuals from regularised negative binomial regression using the modified SCTransform v2 approach. Tumours were integrated into the same multidimensional space using these residuals by canonical correlation analysis (CCA). Uniform Manifold Approximation and Projection (UMAP) plots were constructed using the first 50 dimensions, with initial clustering by shared nearest neighbour evaluation followed by modularity optimisation at a resolution of 0.3, as offered by the FindClusters function. First-order clusters were identified by canonical marker expression, with second-order Leucocyte populations similarly identified. Second-order fibroblast and keratinocyte subpopulations were identified by construction of transcription modules based on differentially expressed genes from publicly available CSCC single-cell RNA sequencing datasets [15, 22]. Annotations were assigned based on module scores within subpopulations.

Selected clusters of interest were subclustered using a similar approach, with an adaptive resolution approach to optimise cluster identification amongst variably heterogenous populations, with resolution ranging at 0.1 (fibroblasts, myovascular) to 0.3 (T cells, keratinocytes).

Expression of markers of interest within cell populations was evaluated by both overall transcript count, but (reflecting the low count numbers) also proportion of cells expressing the gene. A gene was considered expressed if its raw count was greater than zero, and greater than the geometric mean expression of a pool of 10 negative control probes within that cell.

### Spatial co-expression analysis

Spatial correlation of gene expression within cell populations was evaluated using InSituCor (insitucor package) [58]. Briefly, an FOV-restricted 50-nearest neighbour network was constructed for each cell based on Euclidean distance, allowing for the construction of an ‘environmental profile’ for each cell. An environmental expression matrix (summating the expression of each gene within each environment) and a confounder matrix (summarizing variables including signal strength, background noise and cell types present) are constructed. These matrices are then correlated to create a conditional correlation matrix. This matrix measures co-expression of genes within the same spatial neighbourhood beyond that explained by cell type and technical factors. Genes exhibited correlated co-expression were constructed into gene modules, with a minimum spatial conditional correlation and module correlation of 0.1 and resolution of 0.02 (default recommendations), and a minimum module size of 5 genes. Each cell was scored with a weighted average of gene expression within each module (based on both a single-cell and environmental level). This enabled calculation of ‘attribution scores’ for each gene within a module by correlating the environment score for each component gene/module with neighbourhood expression of the gene in question by each cell type. Attribution of modules to cell types was undertaken by correlating the maximum value of the above statistic for the cell type in question for each module genes, leading to high scores for cells that contribute heavily to any of a module’s genes.

### Niche construction

Niches were constructed from 50 nearest-neighbour environments, with frequency of each cell type within environments summated and used for dimension reduction. Autocorrelation was mitigated utilising the recently proposed approach of random sampling of neighbours (sampling proportion 0.5) [59]. Clusters were identified in a same manner as transcriptomic clusters, albeit with 10 dimensions used for shared nearest neighbour evaluation. Spatial trajectory analysis to evaluate the progression of cell environments across the TME and tumour bed was undertaken using an adapted Monocle3 approach.

### Ligand-receptor interaction (LRI) analysis

Cell-cell communication pathways were inferred using the SpaTalk package [60]. Cell graph networks were constructed for individual hub cells within each niche with quantification of ligand-receptor co-expressed pairs between adjacent sender and receiver cells. This was undertaken at 10 neighbour and 50 neighbour network size, to permit differentiation between short-range (predominantly cell surface) and long-distance (predominantly soluble mediator) communication networks.

Permutation testing filters and scores significantly enriched LRI, generating an intercellular score, prior to scoring of expression of receptor-coupled downstream pathway enrichment of transcription factors based on KEGG and Reactome knowledge bases (intracellular score). Scores are then combined to permit LRI ranking. Final scores, calculated as the square root of the product of the intracellular and intercellular scores, were calculated for each LRI. To address LR signalling promiscuity, the highest scoring LRI was used where a ligand had multiple receptors, and the highest intracellular score to identify the most perturbed signalling pathway. LRI with an adjusted p value <0.05, LRI score >0.9 and LR co-expression ratio >0.1 were retained for downstream analysis, based on distribution of these parameters across niches (Fig. S3).

### *In vitro* experiments

Human Umbilical Vein Endothelial Cells (HUVEC, Lonza, Slough, UK) were cultured at 37 °C and 5% CO2 and were used between the fourth and sixth passage. Cells were grown in Endothelial Cell Growth Basal Medium (Lonza, Slough, UK) and supplemented by Fetal bovine serum (FBS) (titer: 1:50), Bovine Brain Extract (titer: 1:250), and GA-1000 (titer: 1:1000). Recombinant human TGF-β1, TGF-β2, TGF-β3 and IL-1β (Biolegend, San Diego, US) were added to culture at 10ng/mL concentration and incubated for up to five days. Media and cytokines were replenished every 48 hours. HUVEC were surface stained (CD31, CD105, CD90) followed by fixation/permeabilization and staining for intracellular markers (Fibronectin, α-SMA and S100A4 (Fibroblast Specific Protein)). Cells in these assays were acquired immediately on the Attune Nxt flow cytometer v3.2.1 and analyzed with FlowJo v.10.10.0. Expression of markers within live HUVEC (evaluated by exclusion of a viability dye, Live-Dead Aqua (Thermo Fisher) was evaluated by comparison to control HUVEC cultured in media only, harvested simultaneously, and quantified as a ratio of stimulated/control median fluorescence intensity (MFI). Groups and timepoints were compared by Kruskal-Wallis test or repeated measures mixed effect modelling against baseline. HUVEC morphology in all experimental groups was evaluated by brightfield microscopy (EVO M500 Imaging System, Thermo Fisher).

## Statistical Analysis

All statistical analyses were performed using R (version 4.3.2). Where P values were adjusted for multiple testing, the Benjamini-Hochberg approach was used. A P value of less than 0.05 was taken as significant. Continuous data was assumed to be non-parametric; for example, correlations are reported after Spearman’s rank-order testing.

## Abbreviations

AOI: Area of interest
ASDC AXL+SIGLEC6+: dendritic cells
BSB: Bruker Spatial Biology
CSCC: Cutaneous squamous cell carcinoma
FOV: Field of view
ICP: Immunocompetent participant
infCAF: Inflammatory cancer-associated fibroblast
IRR: Immune-rich region
KTR: Kidney transplant recipient
LOQ: Limit of quantification
myoCAF: Myofibroblastic cancer-associated fibroblast
panCK: Pan-cytokeratin
PADOG: Pathway analysis with downweighting of overlapping genes
pDC: Plasmacytoid dendritic cell
TGFB: (TGF-β) Transforming growth factor beta
TME: Tumour microenvironment
Treg: Regulatory T cell
TRR: Tumour-rich region
TSK: Tumour-specific keratinocyte

