## Extended Data Figures for "Localised signalling networks co-opt TGF-β2 to promote an immuno-exclusive mesenchymal niche within human squamous cell carcinoma"

### Extended Data Figure 1

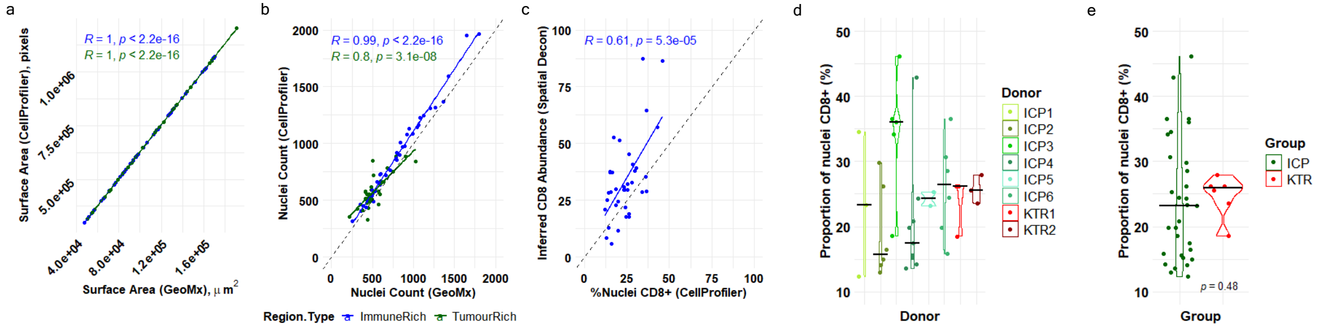


**Extended Data Figure 1: Histological characterisation of AOI.** (A) Cross-platform correlation of surface area and nuclei count (B) of AOI (tumour-rich in green, immune-rich in blue) as calculated by CellProfiler and as measured by GeoMx DSP – correlation (denoted by coloured lines) was evaluated by Spearman rank test; (C) Correlation of proportion of nucleated CD8+ cells amongst all nucleated cells by CellProfiler and inferred CD8 abundance using spatial deconvolution. (D) Proportion of nucleated cells positive for CD8 staining within AOI, stratified by donor. Immunocompetent (ICP) donors are shown in shades of green closed circles, and kidney transplant recipient donors (KTR) in shades of red closed circles. (E) Proportion of nucleated cells positive for CD8 staining within immune-rich AOI, stratified by immunosuppression status (ICP green closed circles, KTR in red closed circles). Difference between groups was evaluated by Kruskal-Wallis test.

### Extended Data Figure 2

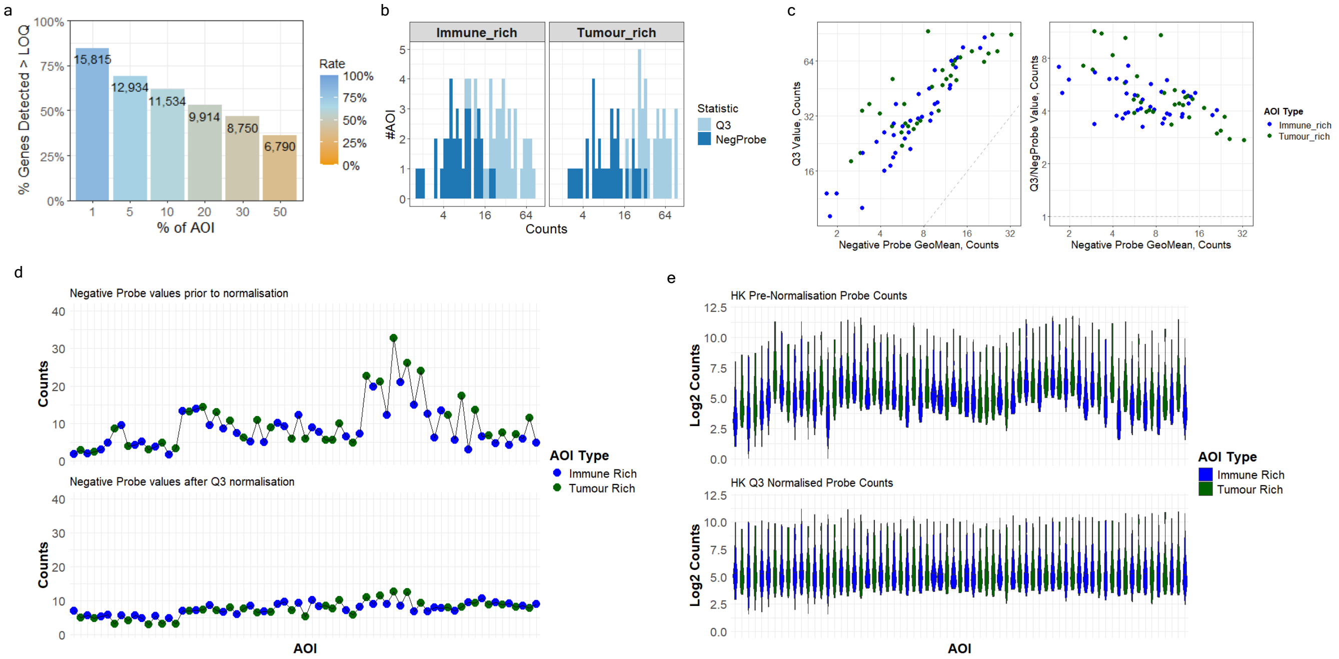


**Extended Data Figure 2: AOI and probe QC.** (A) Boxplot indicating the total number of genes detected above the limit of quantification (LOQ) in different percentages of AOI. For example, 6,790 probes were detected in at least 50% of AOI (right-most bar). A 5% detection rate threshold (yielding 12,934 genes for downstream analysis) was used. (B) Distribution of negative probe and Q3 normalised counts stratified by AOI type. Light blue indicates Q3-normalised probes, dark blue indicates negative control probes. (C) Scatter plot of Q3 normalised counts and negative control counts (left) and ratio of Q3 count/Negative Probe Geometric Means against Negative Probe geometric mean (right) for each AOI, stratified by AOI type. Closed blue circles indicate Immune Rich AOI, green circles indicate Tumour-rich AOI, Grey circles indicate peritumoral normal skin (D) Geometric mean negative probe count prior to (top) and following Q3 (75% percentile) normalisation (bottom), stratified by AOI. Colour indicates AOI type (blue circles = Immune-Rich, green circles = Tumour-rich). (E) Violin plot demonstrating distribution of counts of a panel of 37 commonly used housekeeping genes prior to (top) and following (bottom) Q3 normalisation.

### Extended Data Figure 3

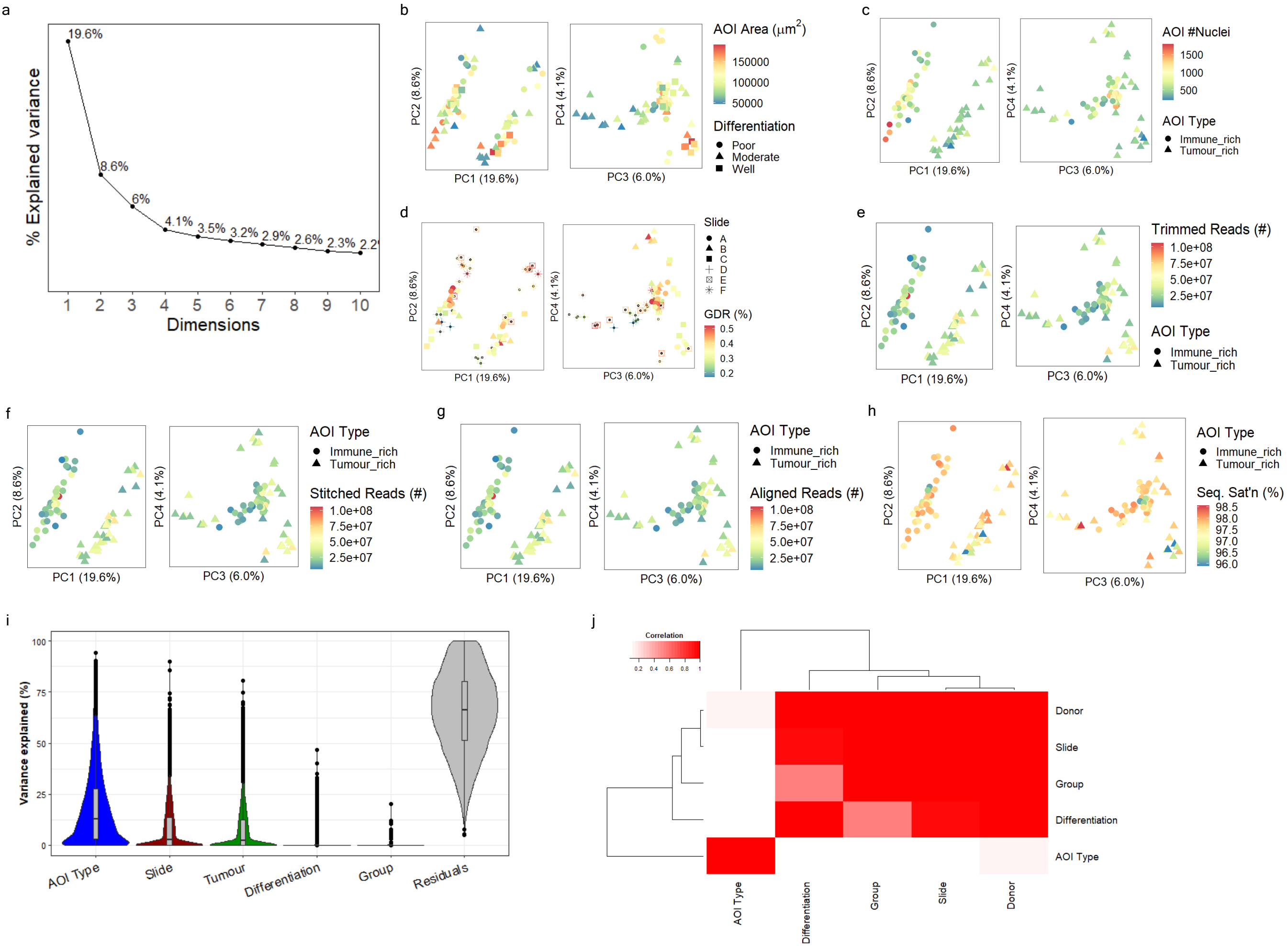

**Extended Data Figure 3: Examination of confounding technical and biological variables by dimensional reduction**. (A) Scree plot of principal components, demonstrating elbow point after 4 dimensions. (B-H) Principal component analysis plot of first four dimensions, annotated for (A) AOI Type / Immunosuppression state (dimensions 3 and 4 only); (B) AOI Area / Tumour Differentiation; (C) Nuclei count within individual AOI; (D) Slide ID / Gene Detection Rate (GDR); (E) Number of trimmed reads within individual AOI; (F) Number of stitched reads within individual AOI; (G) Number of aligned reads within individual AOI; (H) Sequencing saturation within individual AOI; (I) Multivariate linear modelling to evaluate independent contribution of variables to transcriptomic variation. Blue = AOI Type; Red = Slide ID; Green = Donor (Tumour); Yellow = Tumour differentiation; Purple = Immunosuppression Status (Group); Grey = Residual. (J) Canonical correlation heatmap denoting correlation between technical variables. White = low correlation; Red = high correlation.

### Extended Data Figure 4

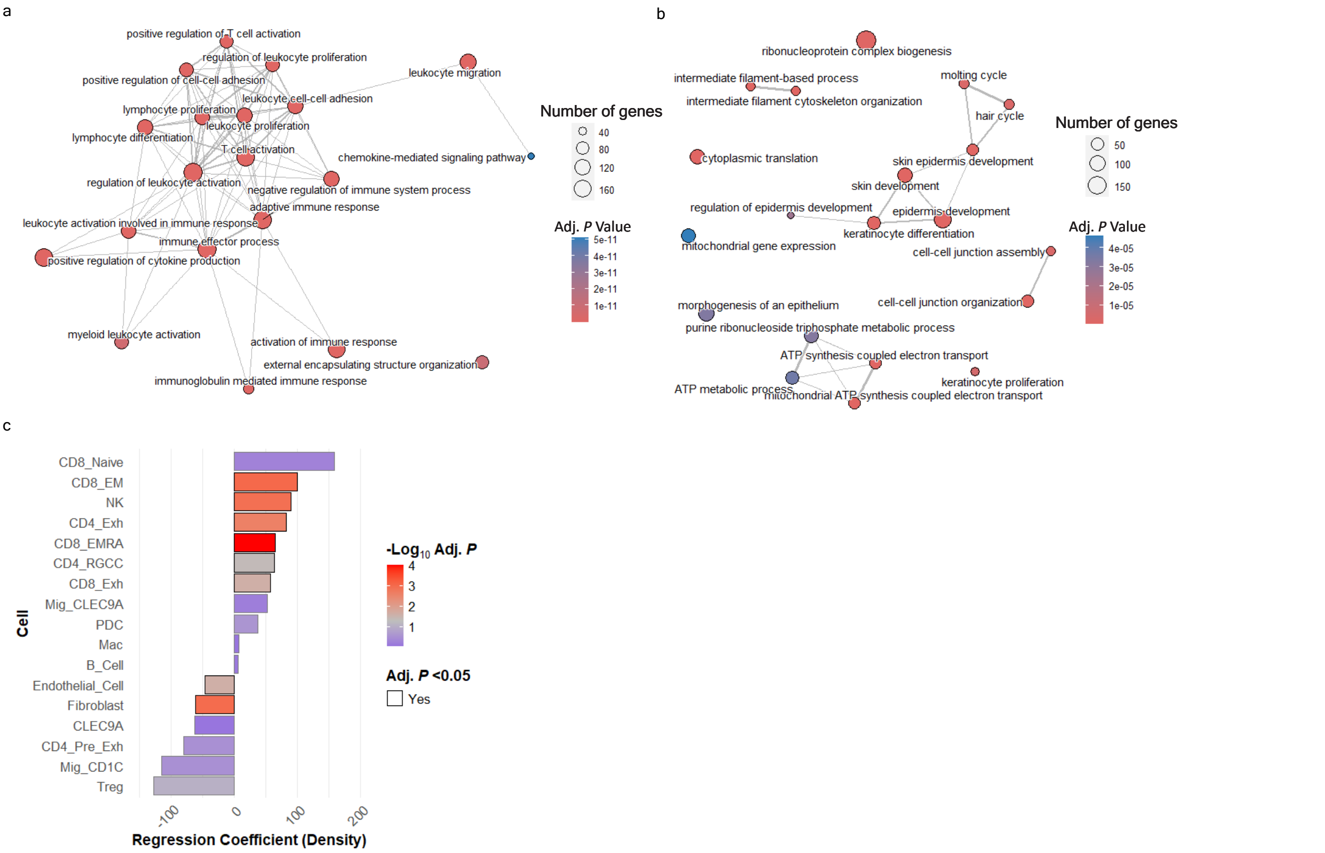


**Extended Data Figure 4: Gene set enrichment and cellular deconvolution analysis.** Enrichment map for top twenty enriched gene sets using Gene Ontology (Biological Processes) knowledgebase, for (A) immune-rich regions and (B) tumour-rich regions. Node size indicates size of gene set, and colour denotes adjusted P value from high (blue) to low (red). (C) Linear modelling of inferred cell abundance and histologically-determined CD8+ T cell density across IRR AOI, controlling for donor. Solid black outline indicates adjusted P value <0.05, with colour denoting adjusted *P* value from high (blue) to low (red), transitioning through grey at adjusted *P* 0.05.

### Extended Data Figure 5

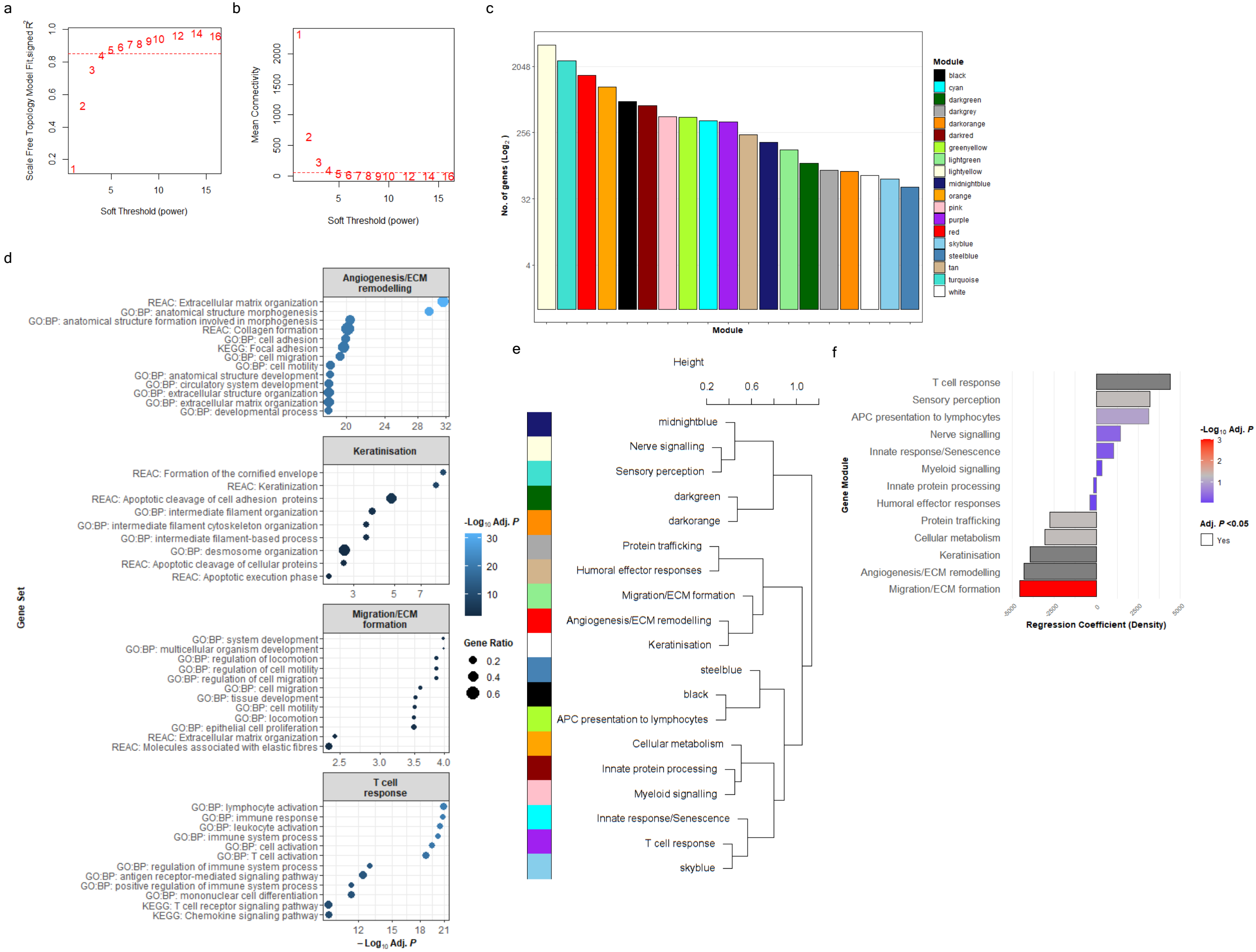


**Extended Data Figure 5: Weighted co-expression gene network analysis of immune-rich regions.** Diagnostic plots showing (A) Scale fit topology and (B) mean connectivity plotted against potential soft threshold values. An $R^2$ of 0.85 and mean connectivity of 50 was set as the threshold for picking the soft threshold for downstream analysis. (C) Barplot denoting final merged module size in descending order. (D) Top enriched gene sets within keratinisation, Migration/ECM formation and T cell response gene networks (a maximum of twelve gene sets are shown for each module, ordered by adjusted P value). (E) Dendrogram indicating clustering of modules and module annotation, using the average linkage between module eigengenes (original module colours are shown on left). (G) Result of mixed effect modelling of histologically determined CD8+ T cell density within AOI, and module eigengene enrichment within the same AOI. Tumour (i.e. donor) was treated as a random effect. *P* values are reported from low (red) to high (blue) transitioning through grey at *P* = 0.05 (adjusted for multiple testing).

### Extended Data Figure 6

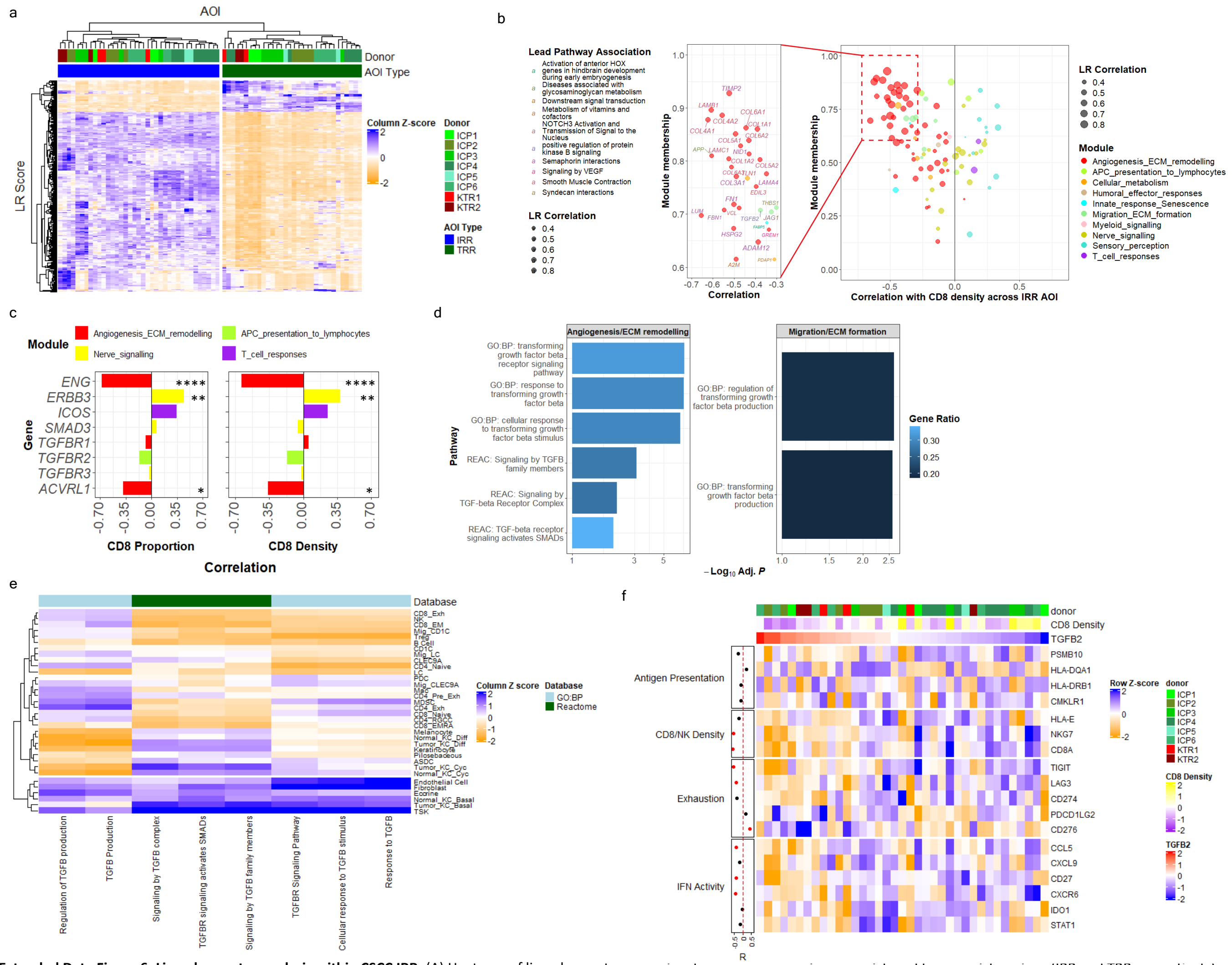

**Extended Data Figure 6: Ligand-receptor analysis within CSCC IRR.** (A) Heatmap of ligand-receptor gene signature scores across immune-rich and tumour-rich regions (IRR and TRR respectively). Column-normalised Z scores are shown (high Z score in blue, low in orange), with hierarchical clustering of LR scores and AOI by complete Euclidean distance. (B) Scatterplot of ligand correlation with histologically-determined CD8+ T cell percentage across AOI, and module membership (dot colour). Inset on left highlights the lead pathway association for those ligands with the strongest negative correlation with CD8 percentage, with label colour denoting lead pathway association and text/dot size indicating LR correlation. (C) TGF-β receptor expression and correlation with CD8 accumulation. Colours indicate co-expression module containing receptor gene (red = Angiogenesis/ECM remodelling; green = APC presentation to lymphocytes; Yellow = nerve signalling; Purple = T cell responses. *$P$ <0.05, ** $P$ <0.01, *** $P$ < 0.001, **** $P$ < 0.0001 by Spearman rank correlation. (D) Location of enriched gene sets with a name containing 'TGF' or 'transforming growth factor' within co-expression modules derived by WGCNA. (E) Enrichment of TGF-related gene sets within cell populations from Ji et al dataset. Enrichment scores are scaled by column. Cell types were clustered by complete Euclidean distance. Normalised gene set enrichment scores are coloured blue (high) to orange (low), with database of geneset origin indicated by blue (Gene Ontology Biological Processes) or green (Reactome). (F) Heatmap illustrating expression of TIS components within IRR from CSCC. IRR (columns) are ordered by local TGFB2 expression (i.e within that region). The dotplot on the left indicates the overall Spearman rank correlation of each gene with TGFB2 expression across all IRR, with those demonstrating signification correlation highlighted in red. Column annotations are by donor, normalised CD8 density (high density in yellow, low density in purple) and normalised TGFB2 expression (high expression in red, low expression in blue).

### Extended Data Figure 7

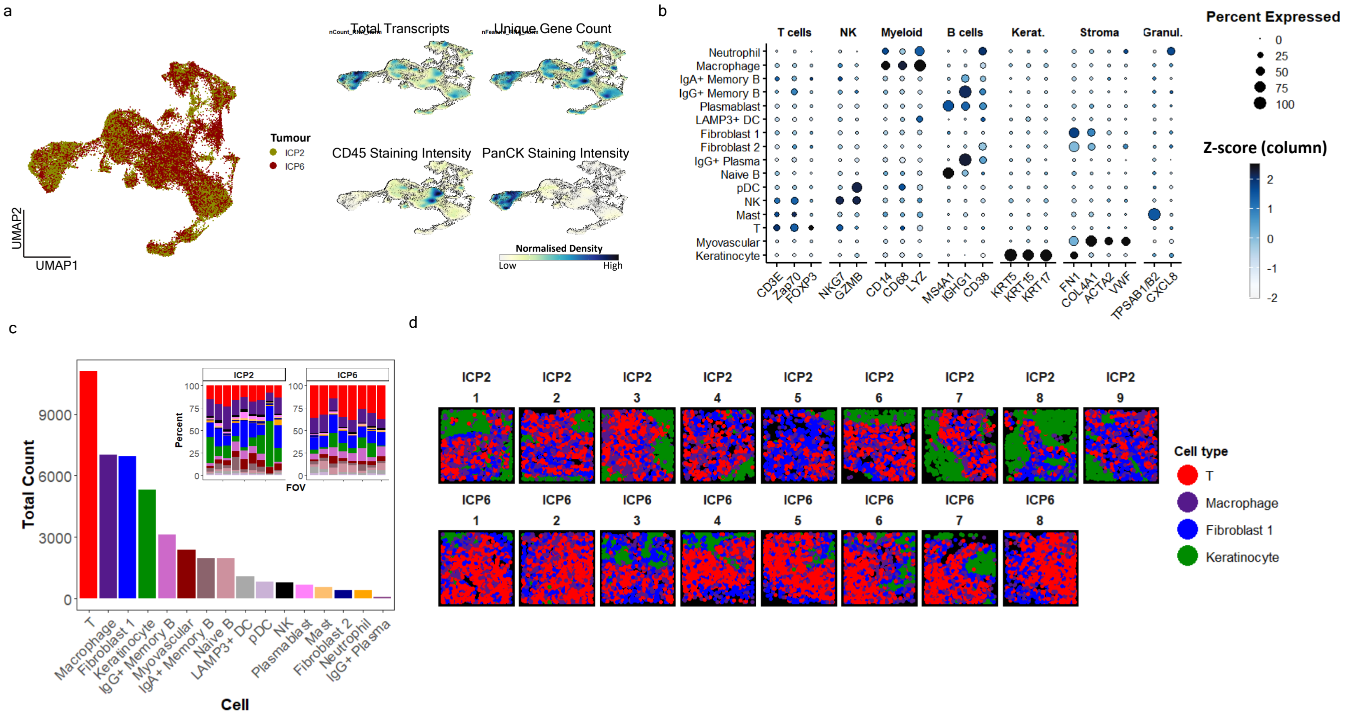


**Extended Data Figure 7: Characteristics of first-order transcriptomic clusters.** (A) UMAP annotated by tumour of origin (left - yellow, ICP2; red; ICP6) and transcriptomic/histological variables (right - normalised density from low (yellow) to high (black)). (B) Expression of canonical markers across clusters - spot size indicates proportion of cluster expressing marker; colour indicates normalised expression level (light blue representing low expression, dark blue representing high expression). (C) Total number of cells detected per cluster across both tumours (inset: proportion of cells per FOV). (D) Location of top 4 most common cell populations in xy space across FOV.

#### Extended Data Figure 8

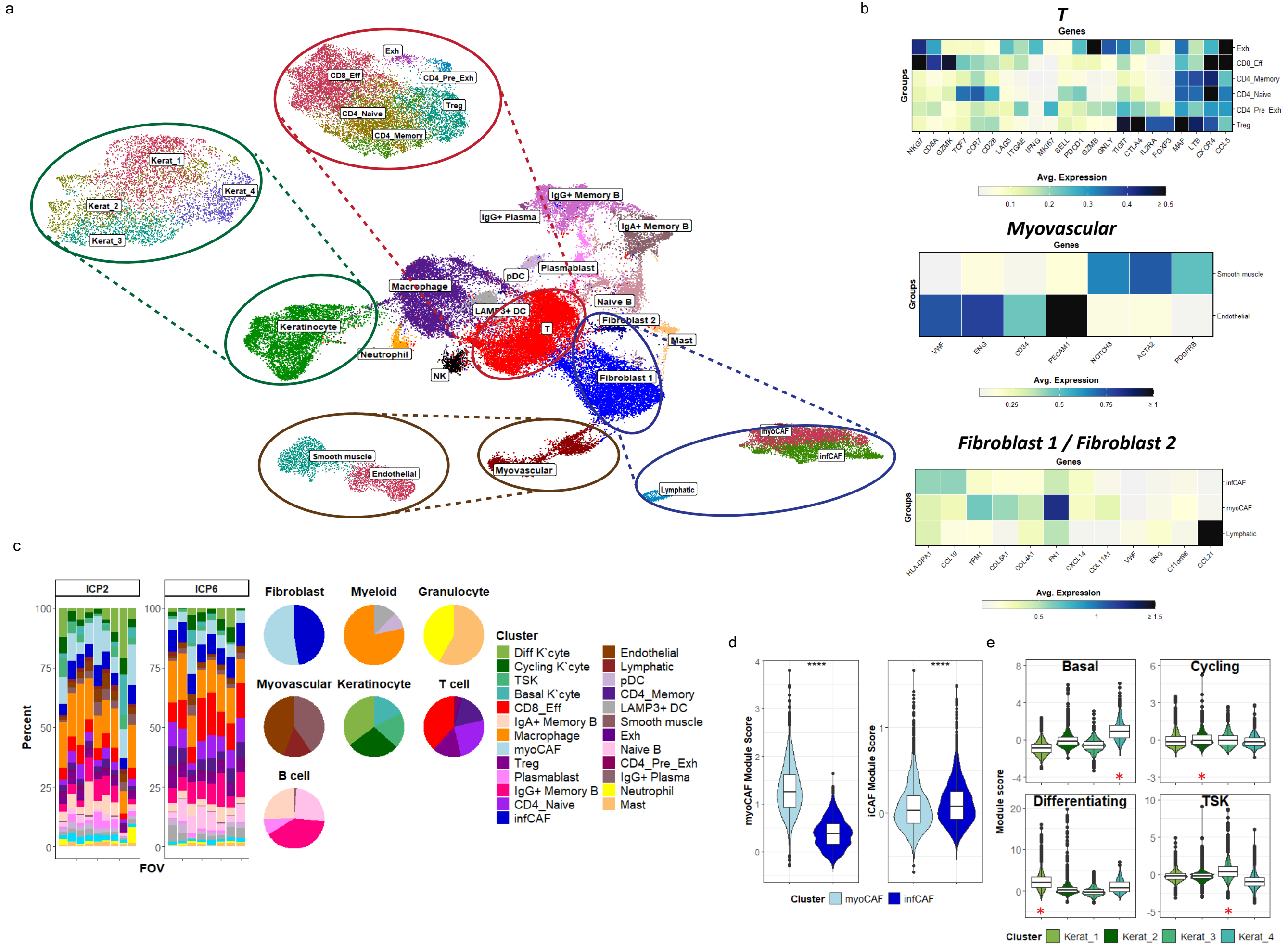


**Extended Data Figure 8: Second order clustering of cell populations.** (A) First-order UMAP, with subclustered UMAPs of T (red ellipses and dashed lines), Keratinocyte (green ellipses and dashed lines), Myovascular (brown ellipses and dashed lines) and Fibroblast1/2 populations (blue ellipses and dashed lines). (B) Heatmaps delineating expression of canonical genes within T, Myovascular and Fibroblast1/2 subclusters. (C) Proportion of second order clusters within FOV (left) and first-order cell clusters (right). (D) Module score for inflammatory and myofibroblast subclusters, using top 30 differentially expressed genes within these cells as previously identified in reference [22]. Light blue violin represents myoCAF, dark blue infCAF. **** *P* <0.0001 by Wilcoxon test. (E) Violin and boxplot of keratinocyte module scores in Kerat_1, Kerat_2, Kerat_3 and Kerat_4 second-order clusters. Module scores were derived from keratinocyte transcriptomic states in Ji et al [15]. Red asterisk indicates the keratinocyte subcluster assigned to the label of the graph.

### Extended Data Figure 9

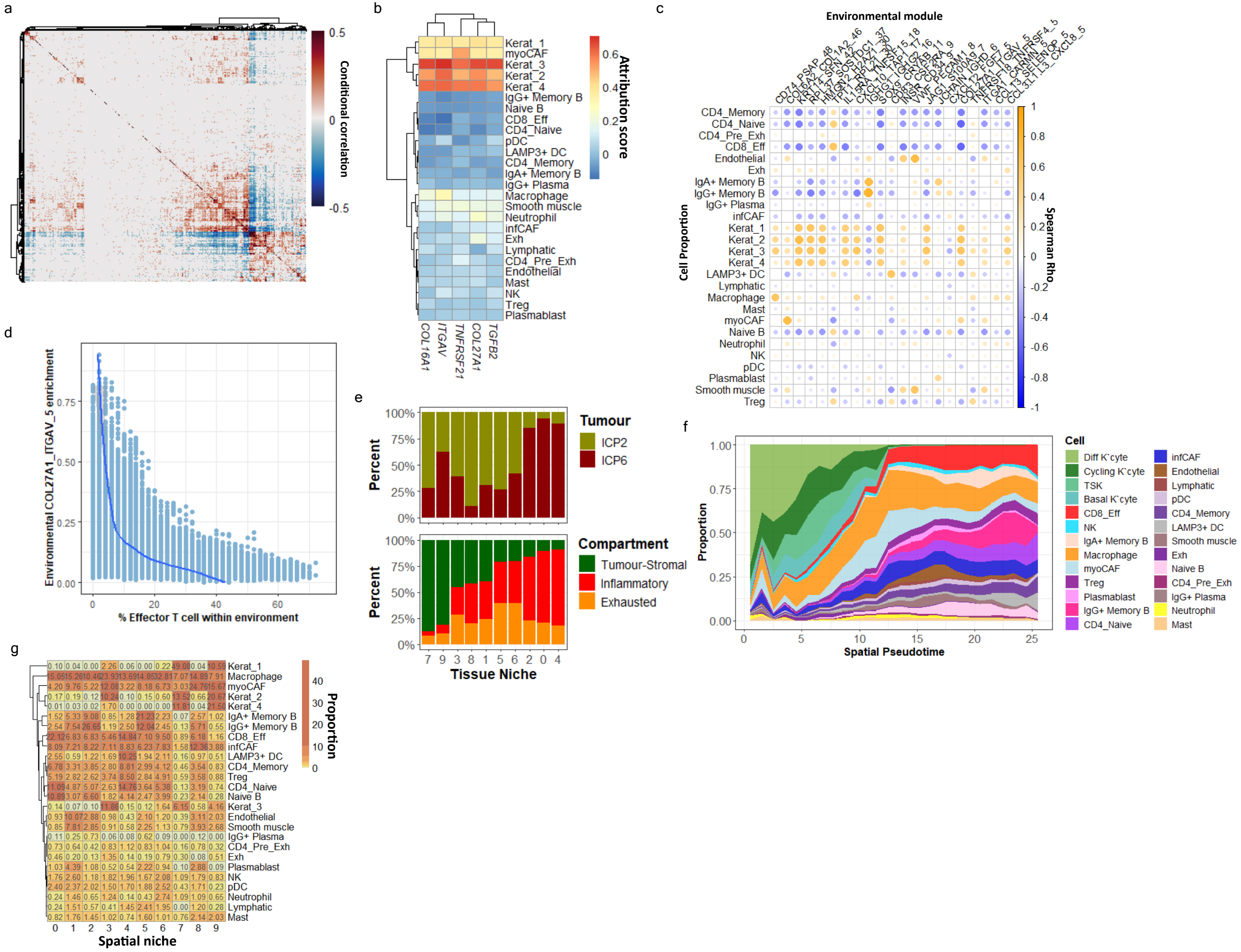


**Extended Data Figure 9: Construction of spatial co-expression gene modules and niches.** (A) Conditional correlation matrix for all 958 genes, showing residual correlation within 50 nearest neighbour (50nn) environments after modelling for local sources of confounding (including signal strength, background noise and cell types present), ranging from blue (low) to red (high). (B) Heatmap of COL27A1_ITGAV_5 module constituent genes and attribution with cell type ranging from blue (low) to red (high). (C) Spearman rank correlation of module enrichment with cell type frequency within 50nn environments, ranging from blue (negative correlation) to orange (positive correlation). (D) Scatterplot of 50 nn environmental COL27A1_ITGAV_5 enrichment and effector T cell proportion (summed proportion of CD8_Eff, CD4_Naive and CD4_Mem). (E) Spatial niche composition stratified by tumour of origin (top, yellow = ICP2, red = ICP6) and cell compartment (bottom, orange = exhausted, green = tumour-stromal, red = inflammatory). (F) Cell type proportions within spatial niches (low proportion = yellow, high proportion = dark orange). (G) Heatmap delineating proportion of cell types within each spatial niche. Low proportion = white/pale yellow, high proportion - orange.

### Extended Data Figure 10

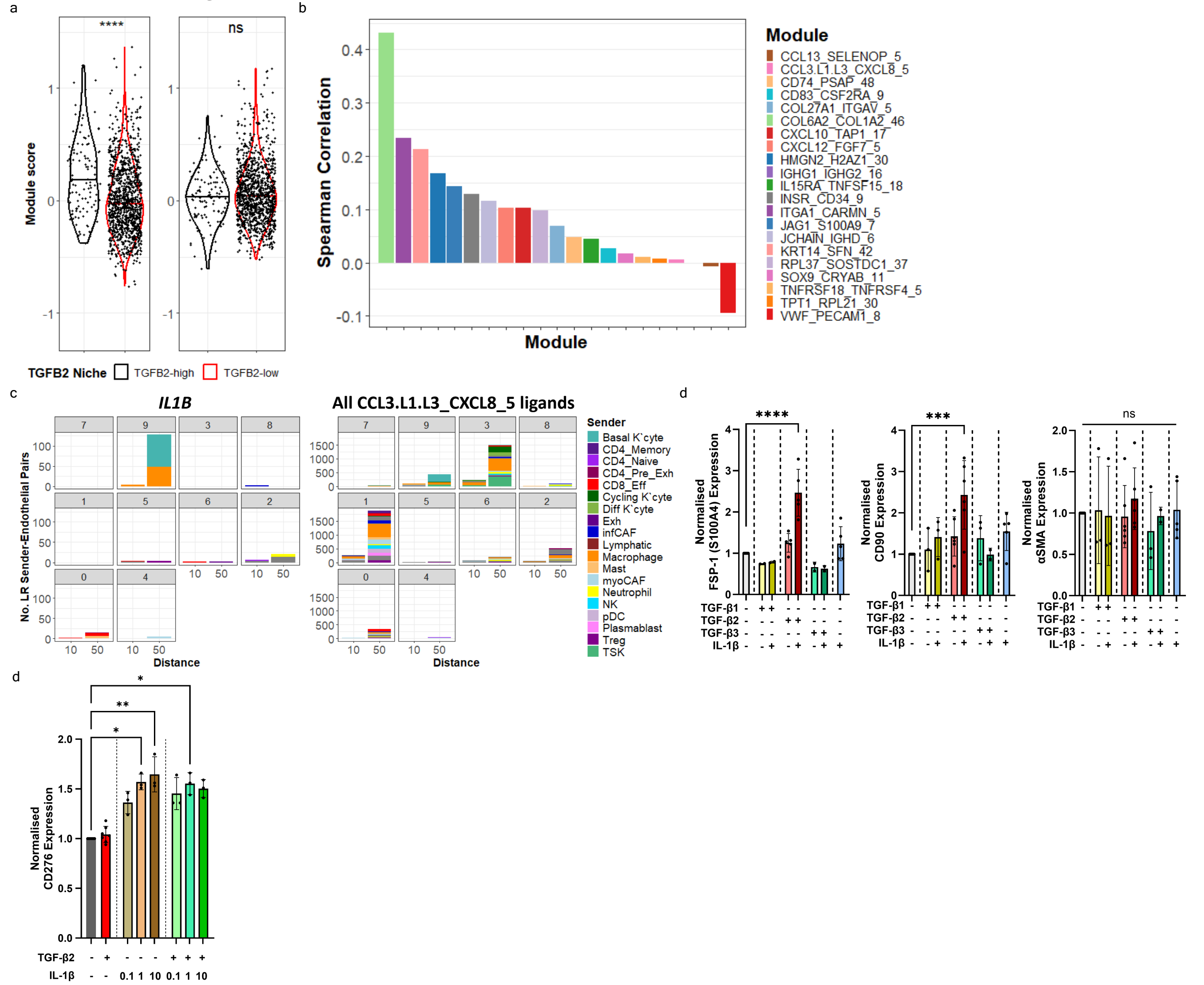


**Extended Data Figure 10: The effect of TGF-β2 and IL-1β upon endothelial cells.** (A) Enrichment of myoCAF and infCAF gene modules within TGFB2-high and TGFB2-low niches. *****P* < 0.0001, ****P* < 0.001 by Kruskal-Wallis test. (B) Spearman rank correlation of myoCAF gene module enrichment with spatial modules enrichment within endothelial cells. (C) Frequency and sender cell of *IL1B* (left) and all CCL3.L1.L3_CXCL8_5 ligands (*CCL3L1/L3*, *CXCL8*, *IL1B* and *CCL4L1/L2* (right) ligand-receptor interaction with endothelial cells by spatial niche at 10- and 50- nearest neighbour resolution. Niches are ordered by pseudotime (reflecting increasing distance from the tumour bed). (D) Expression of CD276 (B7-H3) on HUVEC after 5 days of incubation with the indicated cytokines (IL-1β concentrations are provided in ng/mL, TGF-β2 where used, was at 1ng/mL, with a minimum of three replicates per condition). 'ns' not significant; **P* < 0.05, ***P*<0.01, ****P*<0.001, *****P*<0.0001 by ANOVA with post-hoc Šídák's multiple comparisons test against control media. (D) Expression of S100A4 (left), CD90 (middle) and αSMA (right) in HUVEC after five days culture with specified cytokines (where used, cytokines were added at 1ng/mL). (E) Expression of CD276 in HUVEC after five days culture with specified cytokines (cytokine concentrations are in ng/mL; TGF-β2 was used at 1ng/mL). Only significant differences are shown for S100A4, CD276 and CD90 plots. 'ns' not significant, ****P*<0.001, *****P*<0.0001 by ANOVA with post-hoc Šídák's multiple comparisons test against control media.
