## Supplementary Figures for "Localised signalling networks co-opt TGF-β2 to promote an immuno-exclusive mesenchymal niche within human squamous cell carcinoma"

**KTR1**

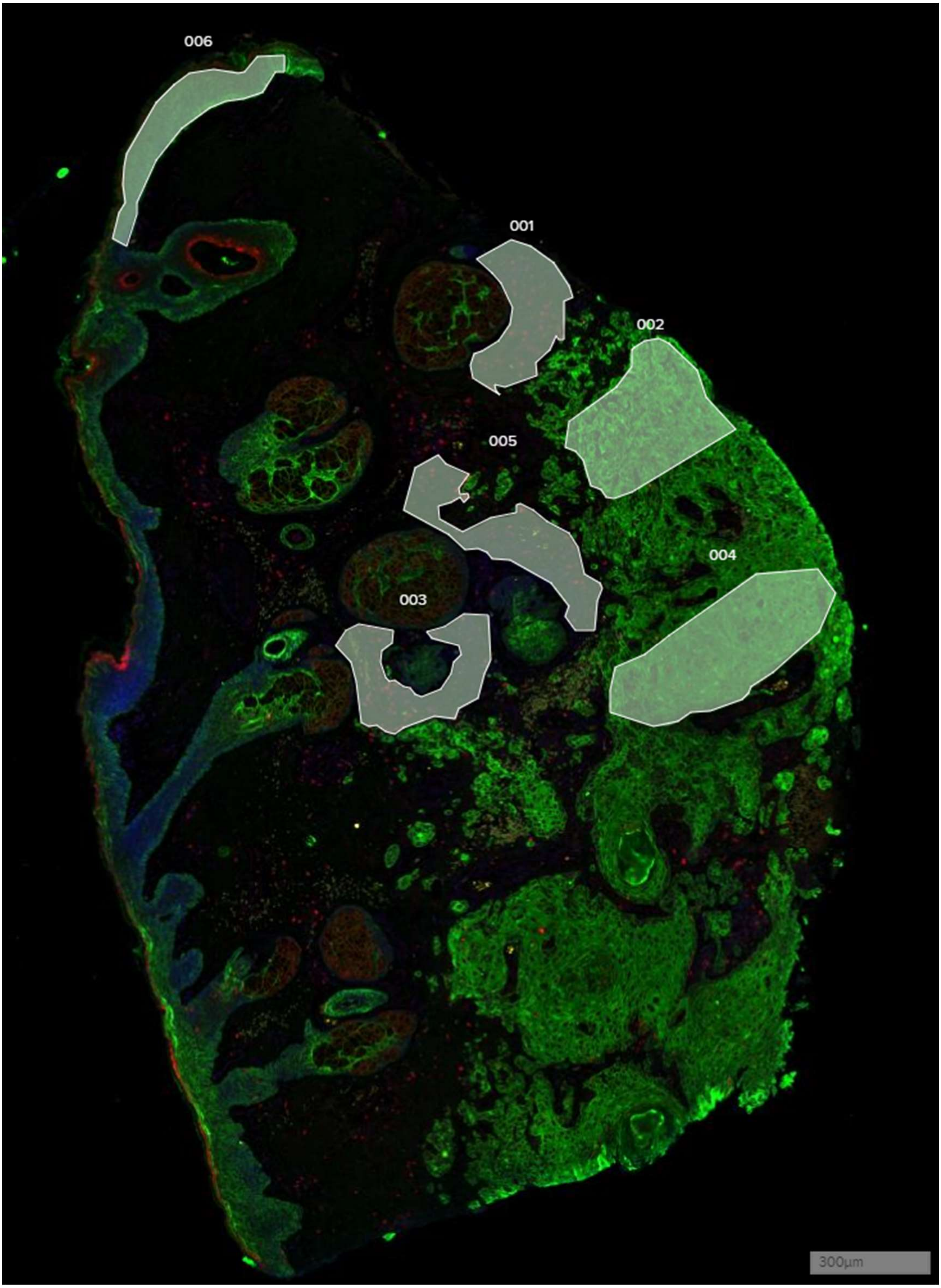

**KTR2**

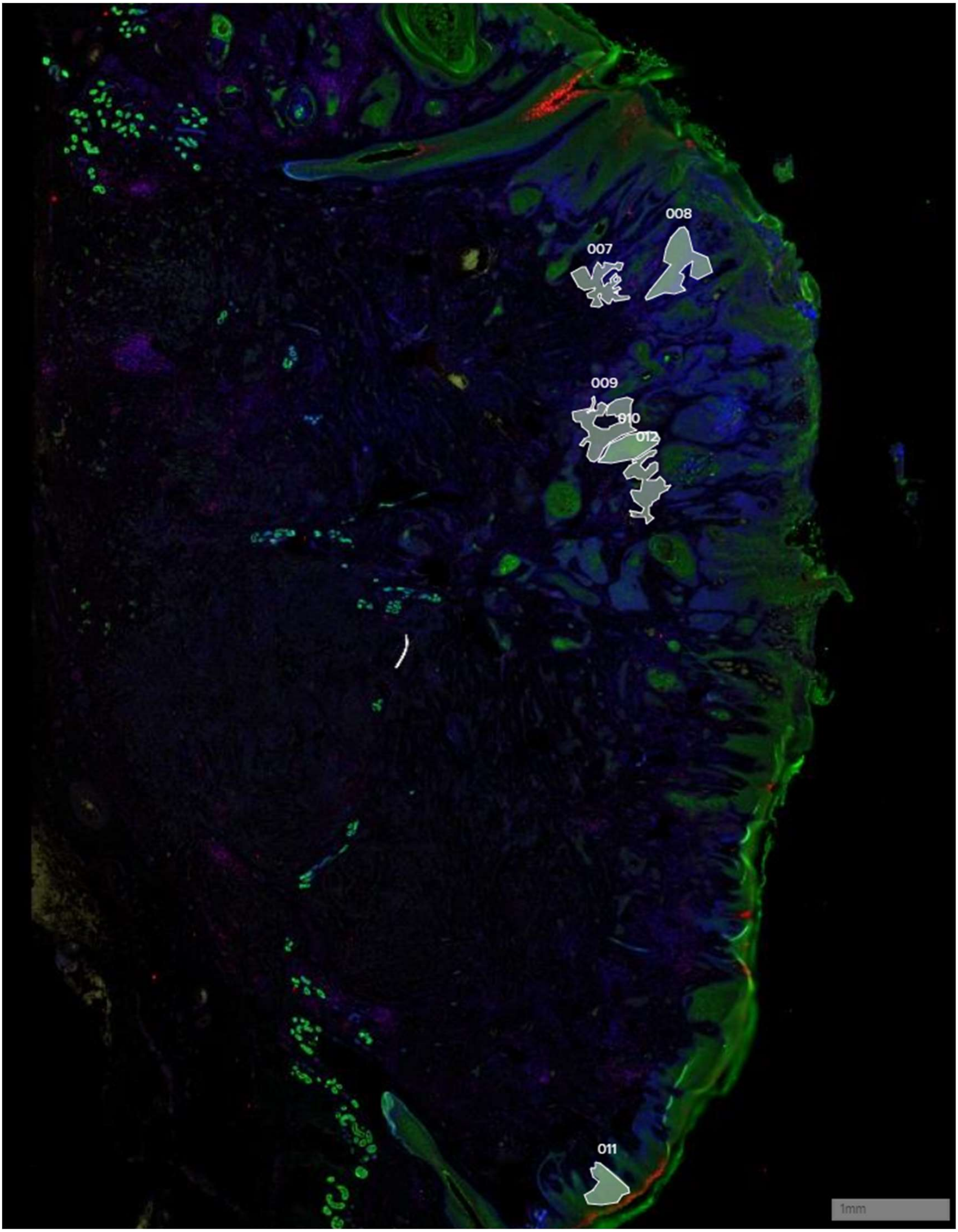

**ICP1**

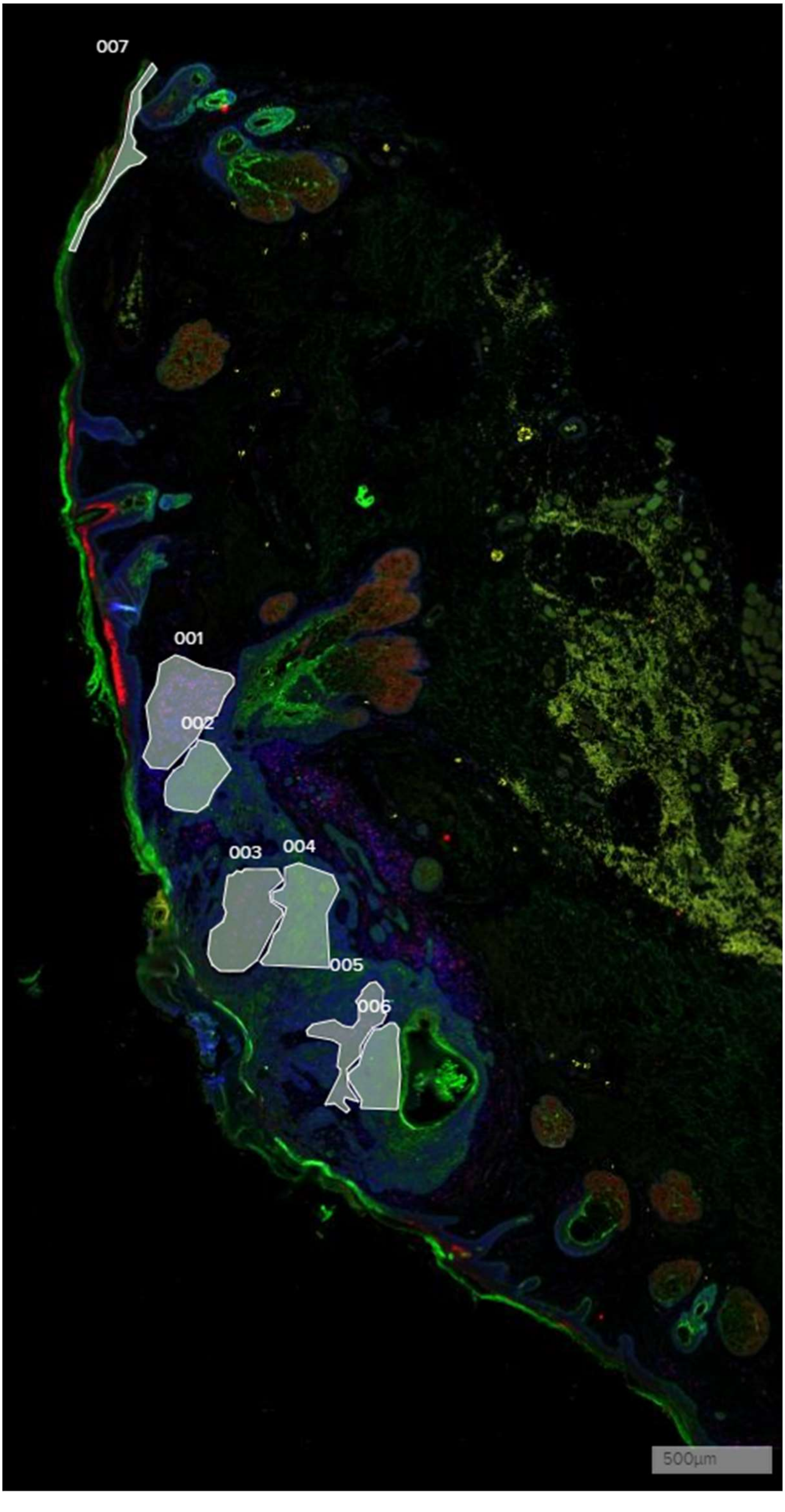

**ICP2**

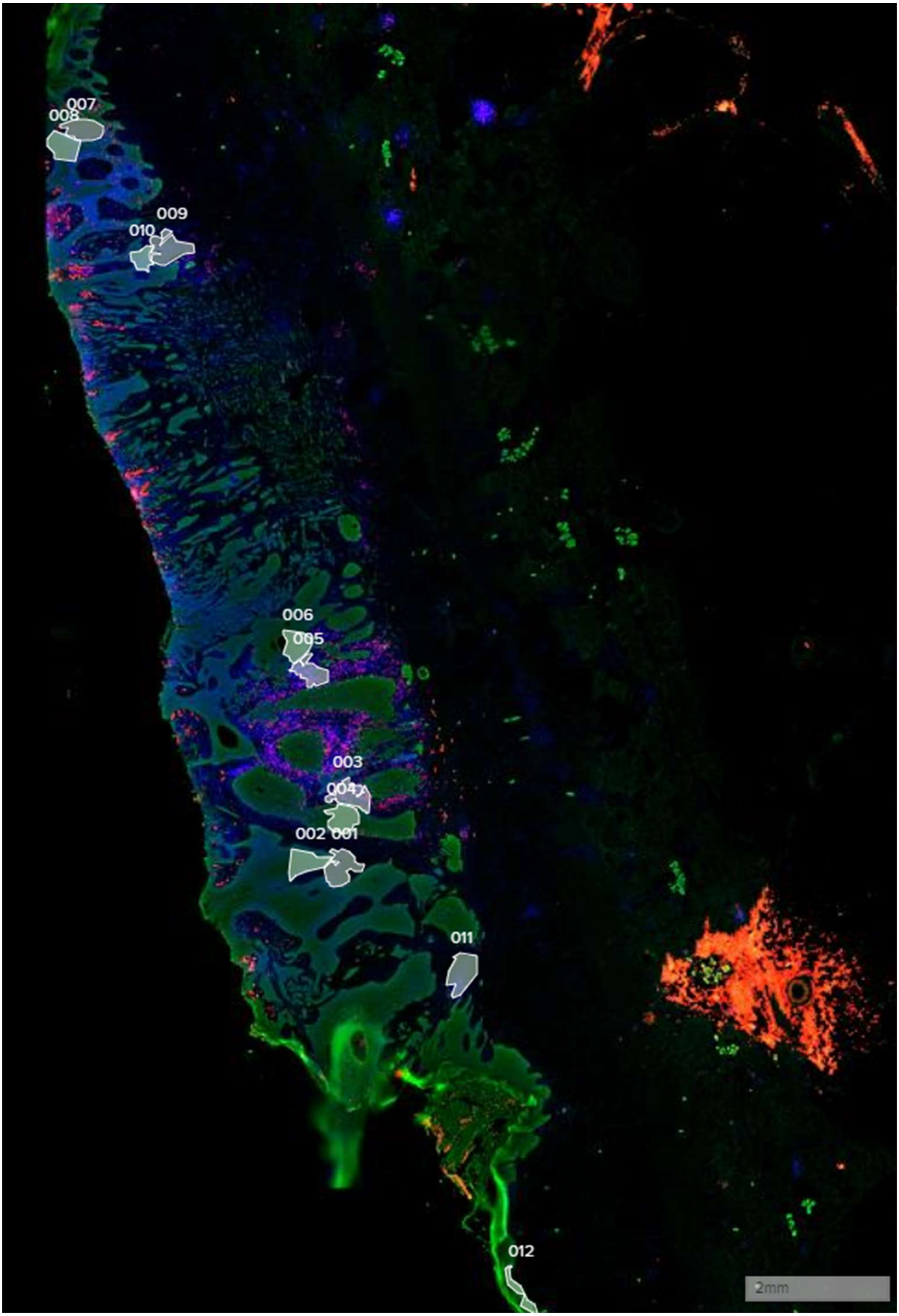

**ICP3**

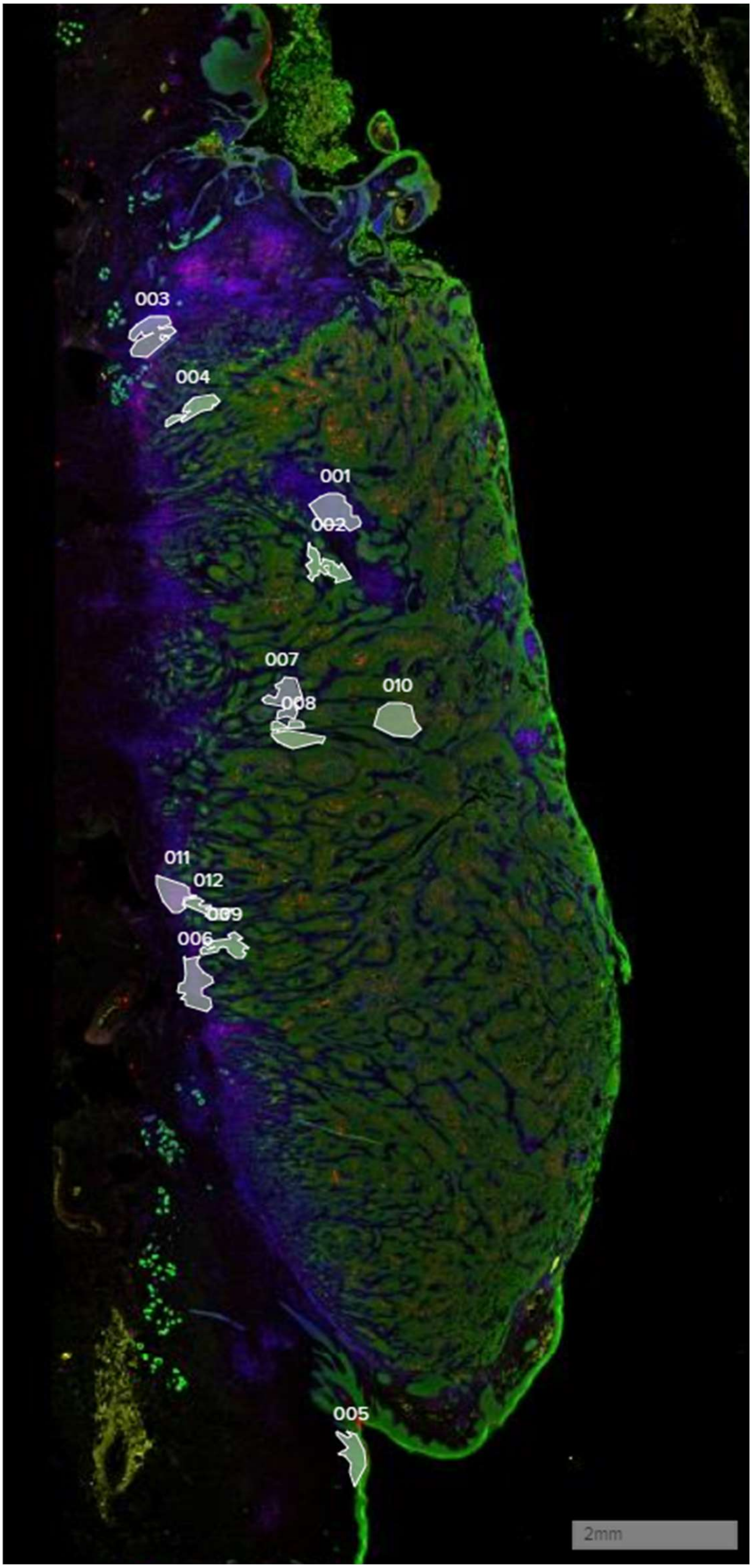

### ICP4

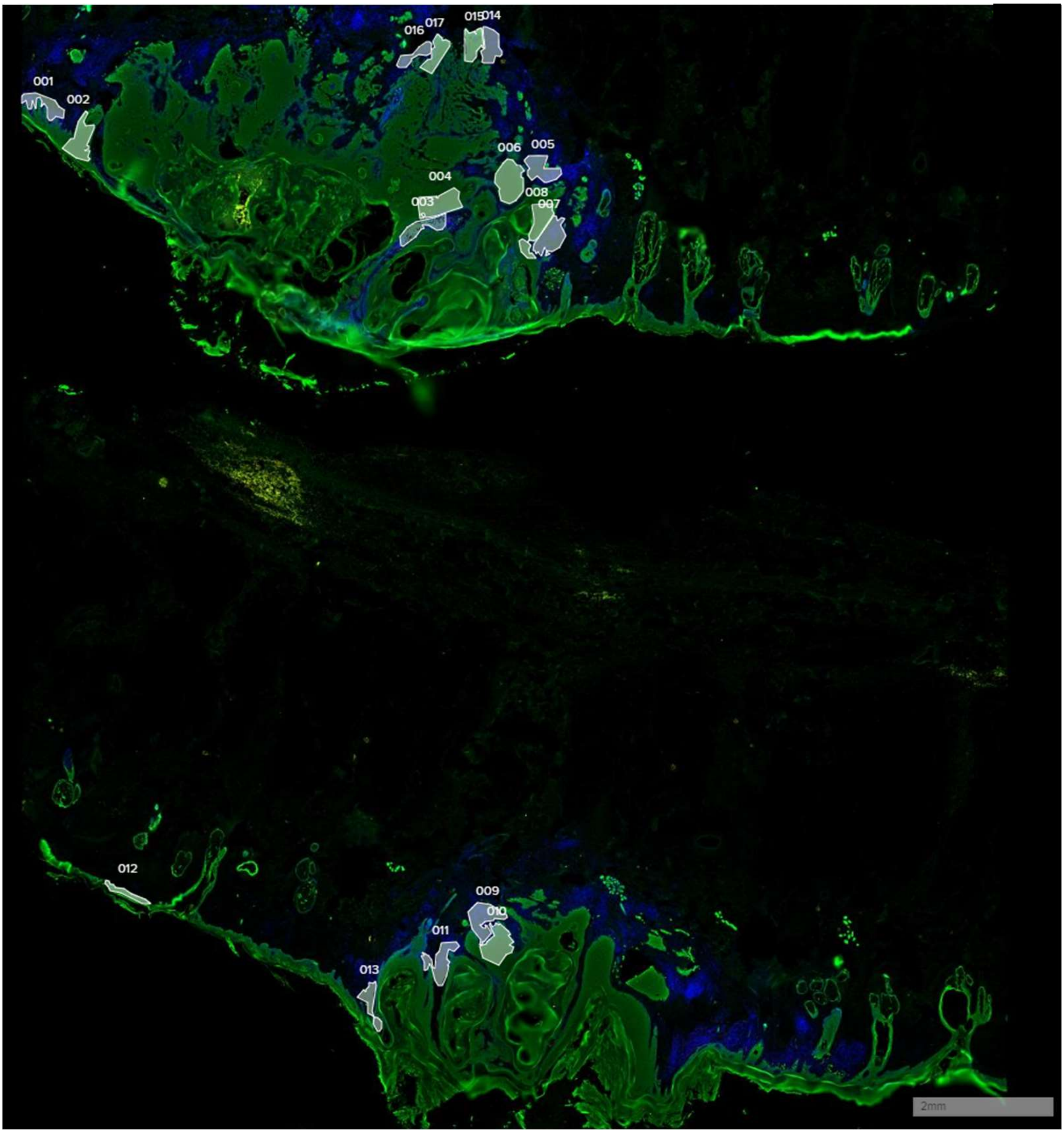
2mm

**ICP5**

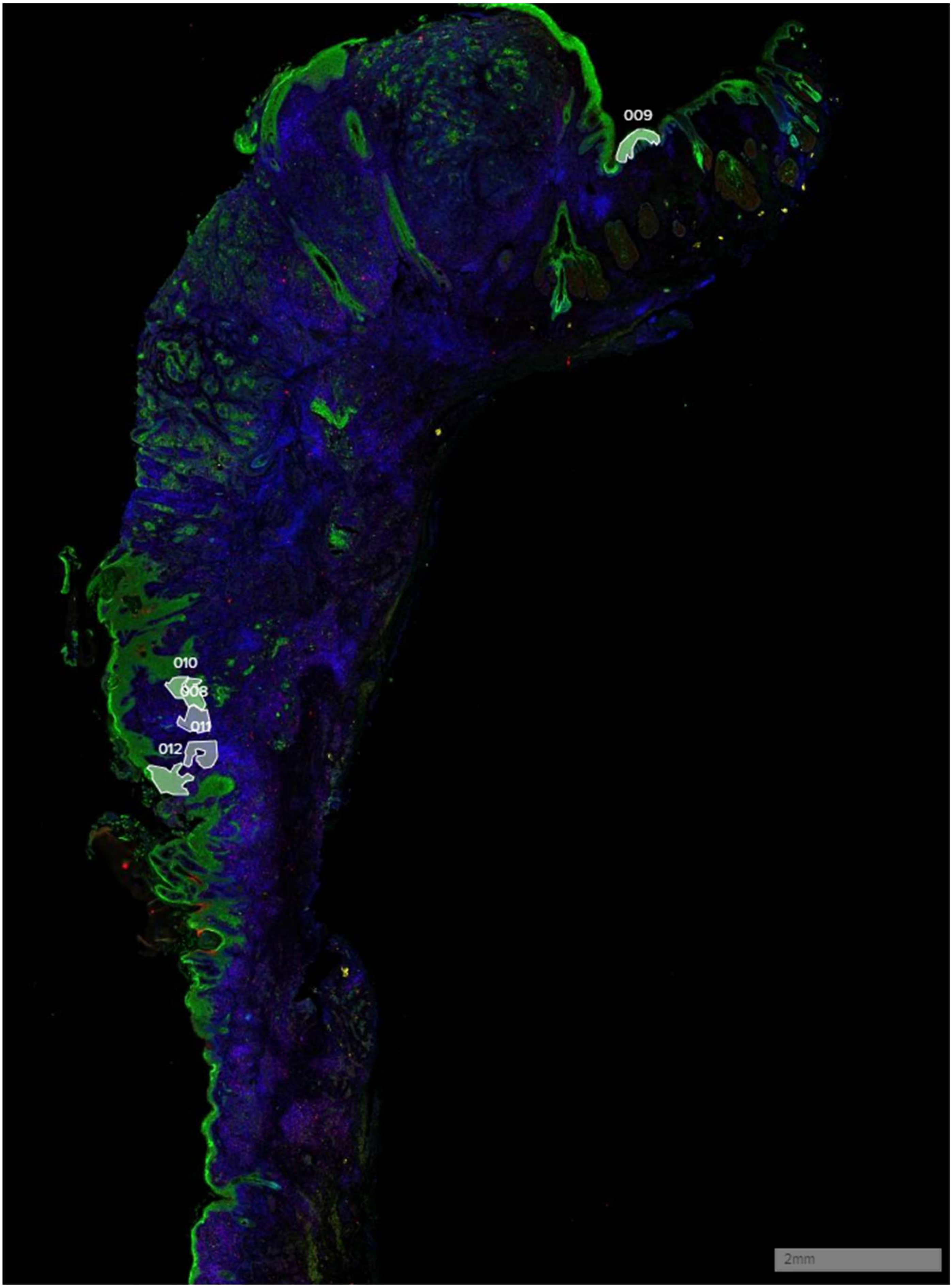

**ICP6**

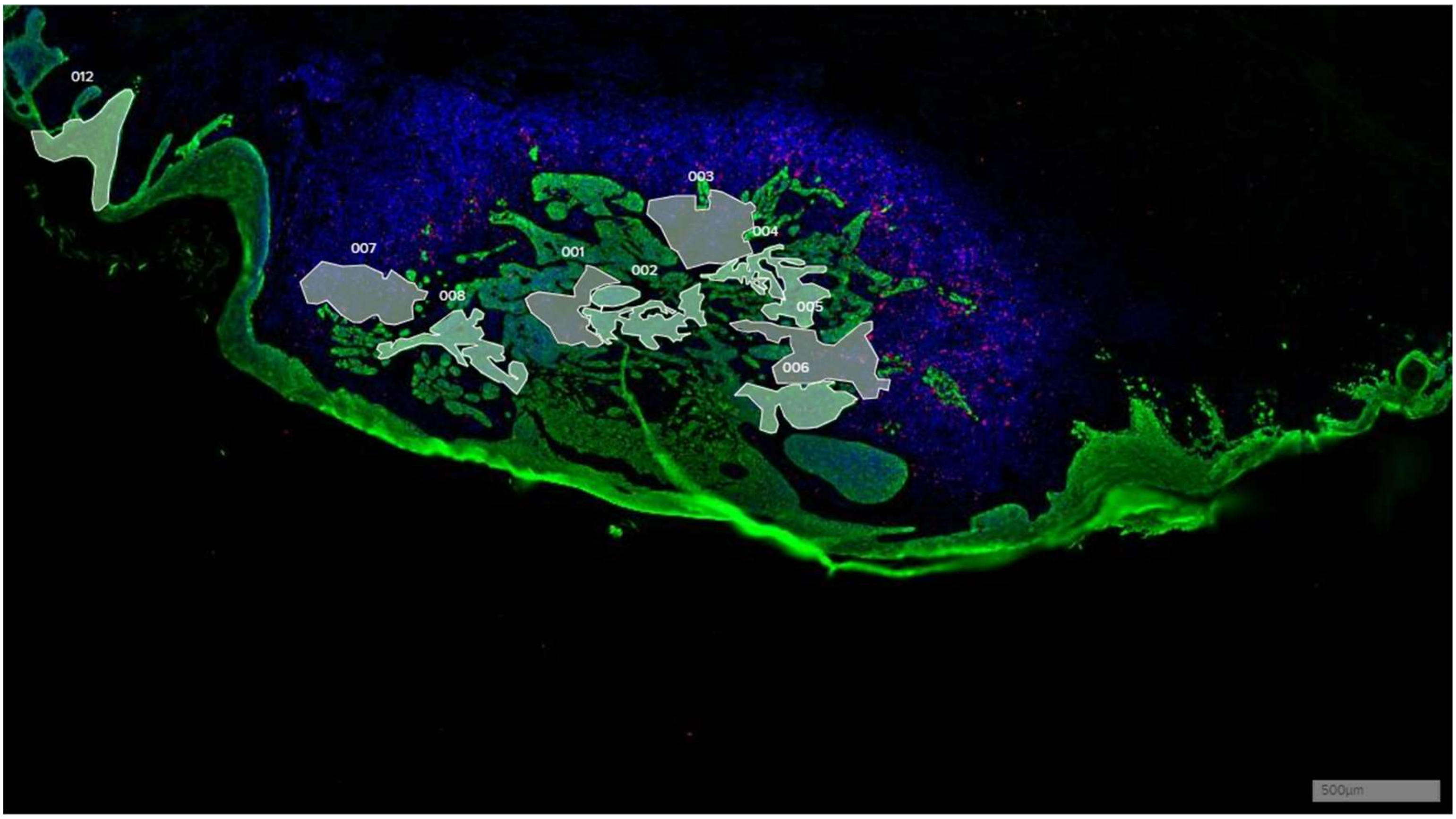

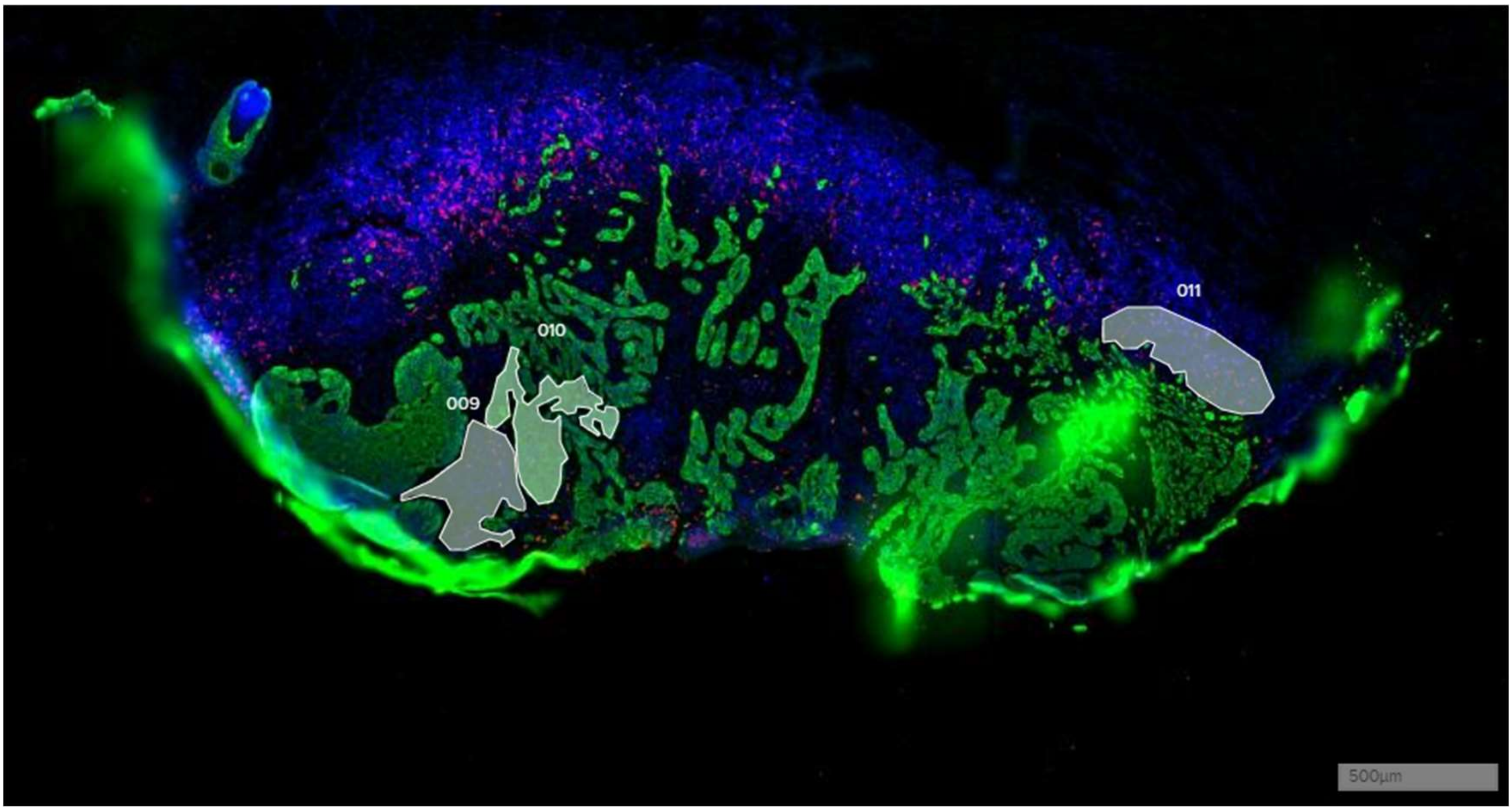

Figure S1: Low power immunofluorescent imaging and designation of CSCC used for GeoMx DSP analysis. ICP6 represents serial sections mounted on the same slide for analysis. White shaded polygons represent areas of interest (AOI) selected for transcriptomic analysis. Blue represents SYTO 13 (nuclear) staining; Green pan-cytokeratin staining; red CD8A staining.

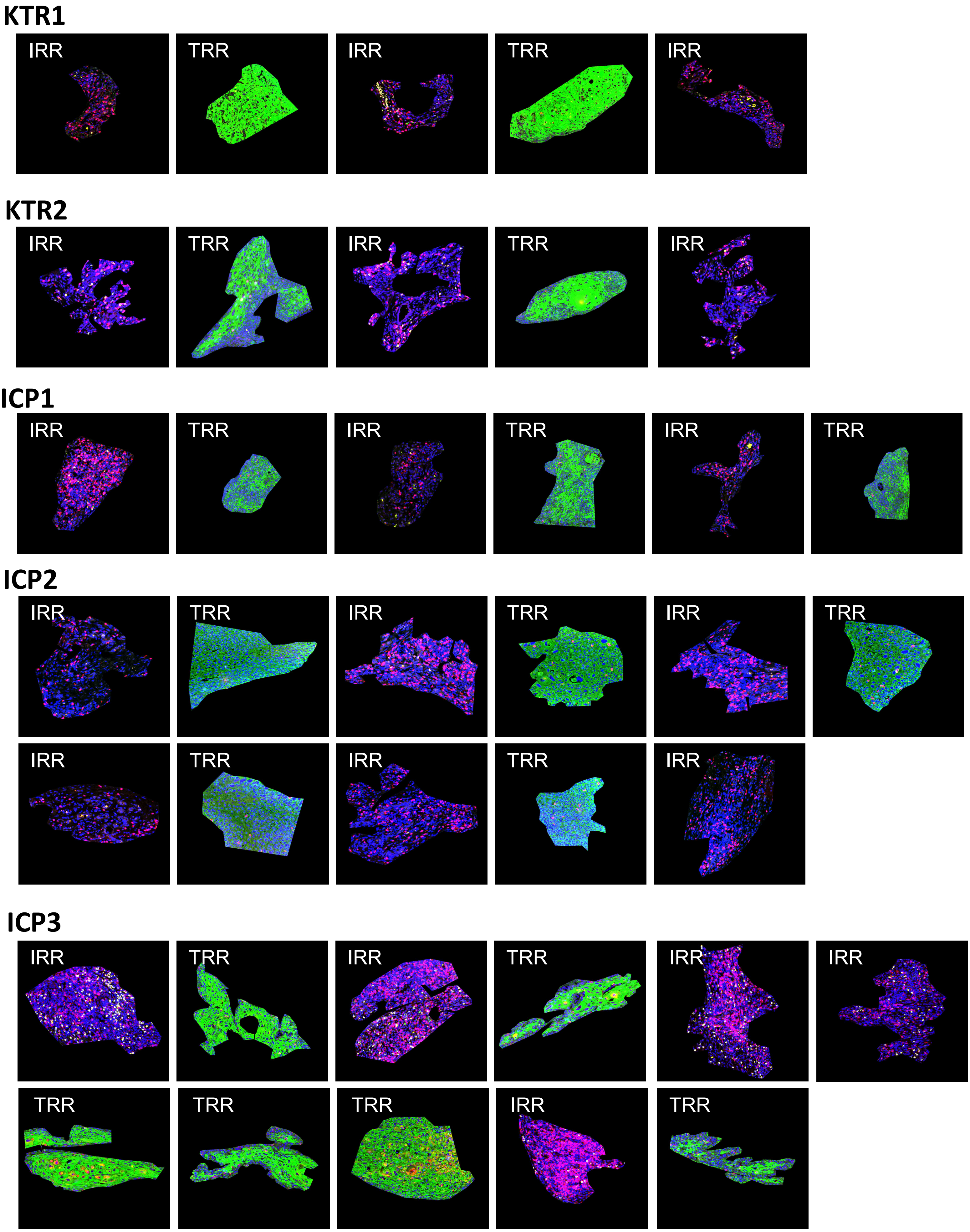
KTR1
IRR
TRR
IRR
TRR
IRR
KTR2
IRR
TRR
IRR
TRR
IRR
ICP1
IRR
TRR
IRR
TRR
IRR
TRR
ICP2
IRR
TRR
IRR
TRR
IRR
TRR
IRR
TRR
IRR
TRR
IRR
ICP3
IRR
TRR
IRR
TRR
IRR
IRR
TRR
TRR
TRR
IRR
TRR

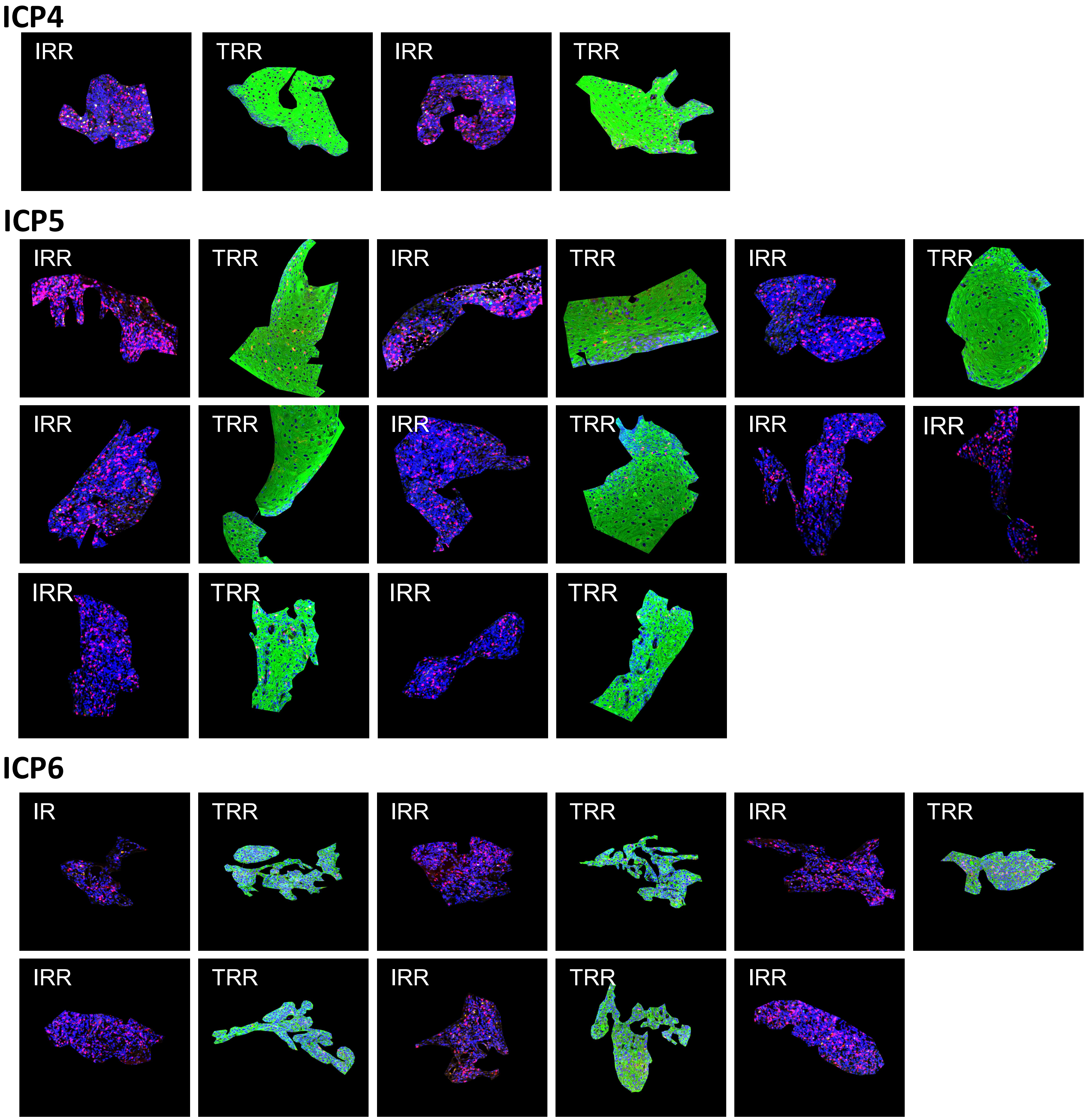

Figure S2: High power immunofluorescent staining and designation of areas of interest (AOI) transcriptomically interrogated by GeoMx. Blue represents SYTO 13 (nuclear) staining; Green pan-cytokeratin staining; red CD8A staining. TRR, tumour-rich region; IRR, immune-rich region.

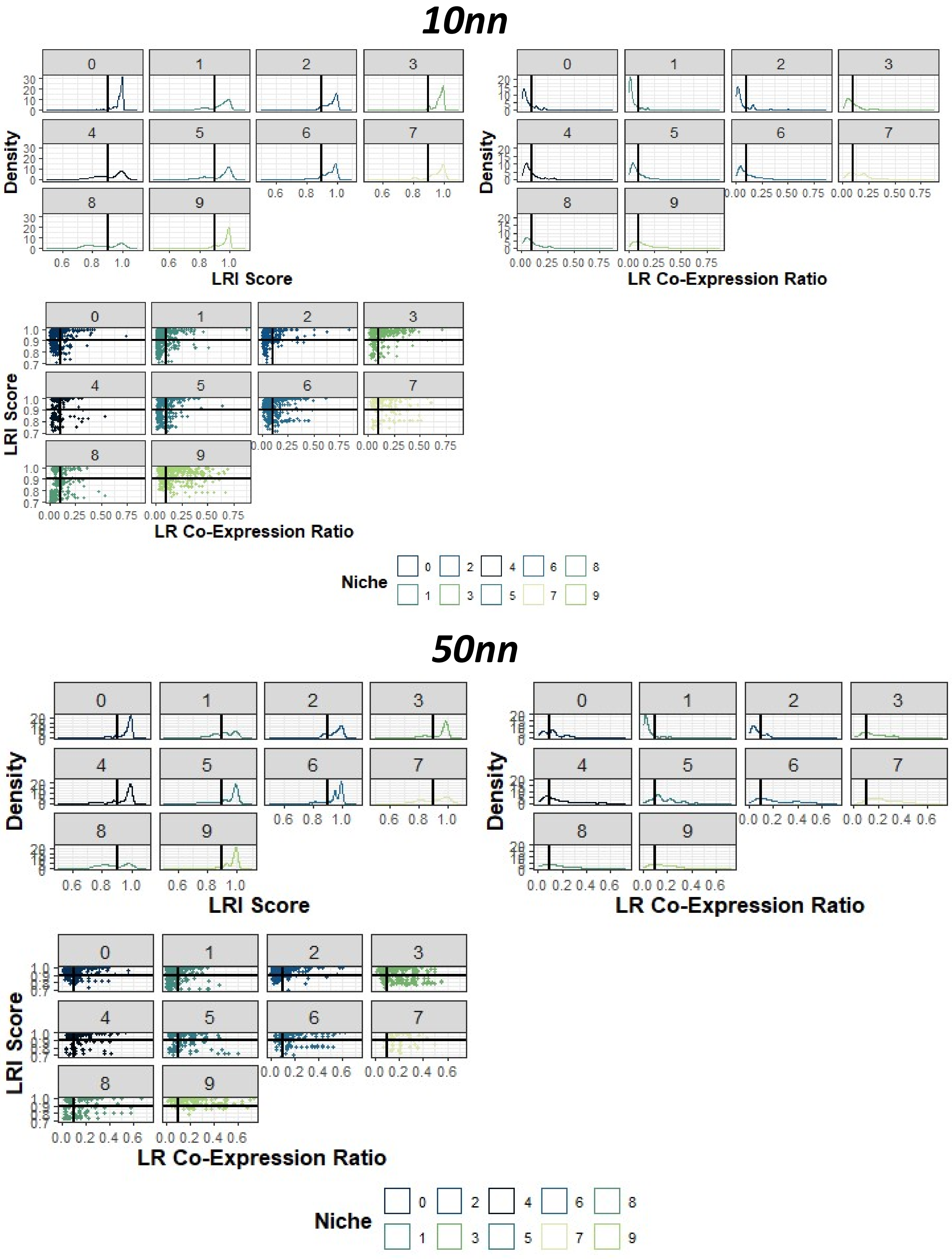

Figure S3: Distribution of ligand-receptor interaction scores and ligand-receptor co-expression ratio across spatial niches at 10 nearest neighbour (top) and 50 nearest neighbour (bottom) resolution. Clockwise from top left – histogram of ligand-receptor interaction scores by spatial niche; histogram of ligand-receptor co-expression ratios by spatial niche; scatter plot of LRI score versus LR co-expression ratio. Black lines indicate thresholds applied for filtering.
